# Brain Damage During New-Onset Refractory Status Epilepticus

**DOI:** 10.1101/2025.08.05.25332982

**Authors:** Cyril F Simmen, Miranda Stattmann, Claudio Togni, Amanda Eisele, Tibor Hortobágyi, Kai Michael Schubert, Andreas Albert Braun, Francesca Casagrande, Carolina Ferreira Atuesta, Niels Briel, Anton Schmick, Marina Herwerth, Anna Lasne, Nadia Mock, Sarah Appenzeller, Sandra Loosli, Rebecca Liu, John S Duncan, Sanjeev Rajakulendran, Matthew C Walker, Aidan Neligan, Jennifer Kilmer, Ferran Prados Carrasco, Stefan Kunst, Nicholas Fearns, Konstantinos Dimitriadis, Moritz Luigi Schmidbauer, Urs Fisch, Raoul Sutter, Stephan Rüegg, Pilar Bosque Varela, Giorgi Kuchukhidze, Eugen Trinka, Johan Zelano, Sarah Akel, Henrik Zetterberg, Pia De Stefano, Margitta Seeck, Dominique Flügel, Dominik Zieglgänsberger, Frederic L.W.V.J. Schaper, Joseph I. Turner, Patrick Roth, Markus Florian Oertel, Nathalie Nierobisch, Matthias J Koepp, Lars Michels, Nicolin Hainc, Giovanna Brandi, Marian Galovic, the Alzheimer’s Disease Neuroimaging Initiative

## Abstract

Status epilepticus (SE) has long been linked to neuronal damage in experimental and animal studies, yet direct human evidence remains scarce. The risk of seizure-induced brain injury is central to the definition, urgency, treatment, and prognosis of SE. We studied 2,055 longitudinal MRI scans from 559 individuals, including 33 patients with new-onset refractory SE (NORSE) across multiple centres, to quantify grey matter volume changes during and after SE. We demonstrate a rapid, widespread, and irreversible decline in grey matter volume during NORSE, exceeding normal aging by 80-fold and Alzheimer’s disease by 20-fold. Fluid biomarkers confirmed marked neurodegeneration during NORSE, correlated with grey matter volume reduction, and returned to low levels after SE. Accelerated atrophy was linked to longer SE duration, poorer long-term outcome, and cryptogenic aetiology. These findings underscore the urgency of treating SE to limit brain damage and provide a framework for evaluating potentially neuroprotective interventions in humans.

---

Since Sir William Gowers noted that *“seizures beget seizures,”*^1^ the question of whether seizures cause brain damage remained a matter of critical importance for both patients and the treating physicians. Evidence for neurodegeneration following brief seizures remains controversial, as findings from animal studies^2–5^ and human neuropathological^6–10^ and neuroimaging^11,12^ analyses have been inconsistent.

In contrast, abnormally prolonged seizures, termed status epilepticus (SE),^13^ have been shown to consistently cause neuronal damage in experimental animal models.^5,14,15^ SE is one of the most common neurological emergencies,^16^ with a mortality rate of 10-20%.^17^ During SE, excessive glutamate release^18,19^ activates NMDA receptors, causing an influx of calcium ions that trigger a neuronal cell death cascade.^20^ Additional mechanisms involving other brain receptors,^21,22^ mitochondrial dysfunction,^23^ production of reactive oxygen species,^24^ and genetic/epigenetic processes^25,26^ may also play a role.

*In vivo* human evidence of SE-induced neuronal loss is confined to case reports^27,28^ and small case series that relied on only rudimentary atrophy metrics.^29,30^ A recent study reported a 16% ventricular expansion after SE with periictal magnetic resonance imaging (MRI) abnormalities, but its interpretation is limited by coarse morphometry and potential confounding from heterogeneous aetiologies and pre-existing structural lesions.^31^ Elevated fluid biomarkers of brain injury^32^ and neuronal^33,34^ or axonal^35,36^ damage have been detected during SE.

New onset refractory SE (NORSE) is a rare and severe form of SE that provides a unique opportunity to study SE-related neurodegeneration. By definition, NORSE occurs in individuals without preexisting epilepsy or other significant neurological disorders, minimizing confounding factors.^37^ Its typically prolonged course^38,39^ allows for serial MRI scanning providing insights into morphometric changes during and after SE. Treatment in intensive care units (ICU) of co-occurring hypotonia, hypoxia, fever, or metabolic disturbances facilitates accounting for confounding systemic factors.^15^ Long-term outcomes, including multidomain cognitive impairment^40,41^ and reports of brain atrophy,^42–44^ suggest significant neurodegeneration in NORSE patients.

We tested the hypothesis that NORSE is associated with significant brain damage by tracking brain atrophy through serial MRI and fluid biomarkers of neurodegeneration. We found substantial, irreversible brain atrophy in subjects with NORSE compared to healthy adults and individuals with other neurological diseases and examined its associations with clinical variables.

## Results

### Longitudinal MRI detects rapid grey matter volume reduction during NORSE

In order to assess the impact of NORSE on brain atrophy and compare these effects with healthy aging and other brain disorders, we assessed a total of 2,055 longitudinal structural MRIs from 559 individuals. The single-centre discovery cohort included 16 NORSE patients with 48 MRIs, while the multi-centre replication cohort consisted of 17 NORSE patients with 42 MRIs from six centres. These were compared against age- and sex-matched healthy controls (n=99; 198 MRIs), patients with epilepsy and brief seizures (i.e. seizures not qualifying as SE; n=115; 230 MRIs), active autoimmune encephalitis without SE (n=31; 75 MRIs), acute subarachnoid haemorrhage without SE requiring ICU care (n=23; 58 MRIs), mild cognitive impairment (n=155; 958 MRIs), and Alzheimer’s disease (n=103; 446 MRIs). Baseline characteristics are detailed in **Extended Data Tables 1 and 2**, and **Supplementary Information**. The discovery cohort was representative of all NORSE cases treated at the University Hospital Zurich, as outlined in **Extended Data Table 3**.

Brain-wide plots of the annualised changes in cortical thickness and subcortical volumes revealed widespread accelerated atrophy in NORSE patients compared to all other groups (**Figure 1**, data on log-scale). The rate of grey matter volume (GMV) reduction (**Figure 2A**) in the NORSE discovery cohort (272.4 ± 57.0 ml/year) was faster than in healthy controls (2.9 ± 1.2 ml/year, p<0.001), epilepsy (3.3 ± 1.9 ml/year, p<0.001), autoimmune encephalitis (4.9 ± 4.6 ml/year, p<0.001), subarachnoid haemorrhage (7.8 ± 3.0 ml/year, p<0.001), mild cognitive impairment (7.1 ± 1.9 ml/year, p<0.001), and Alzheimer’s disease (10.3 ± 2.2 ml/year, p<0.001). Morphometric analysis (**Figure 2B**) showed that accelerated atrophy in NORSE was widespread, affecting bi-hemispheric cortical and subcortical areas.

**Figure 1:**
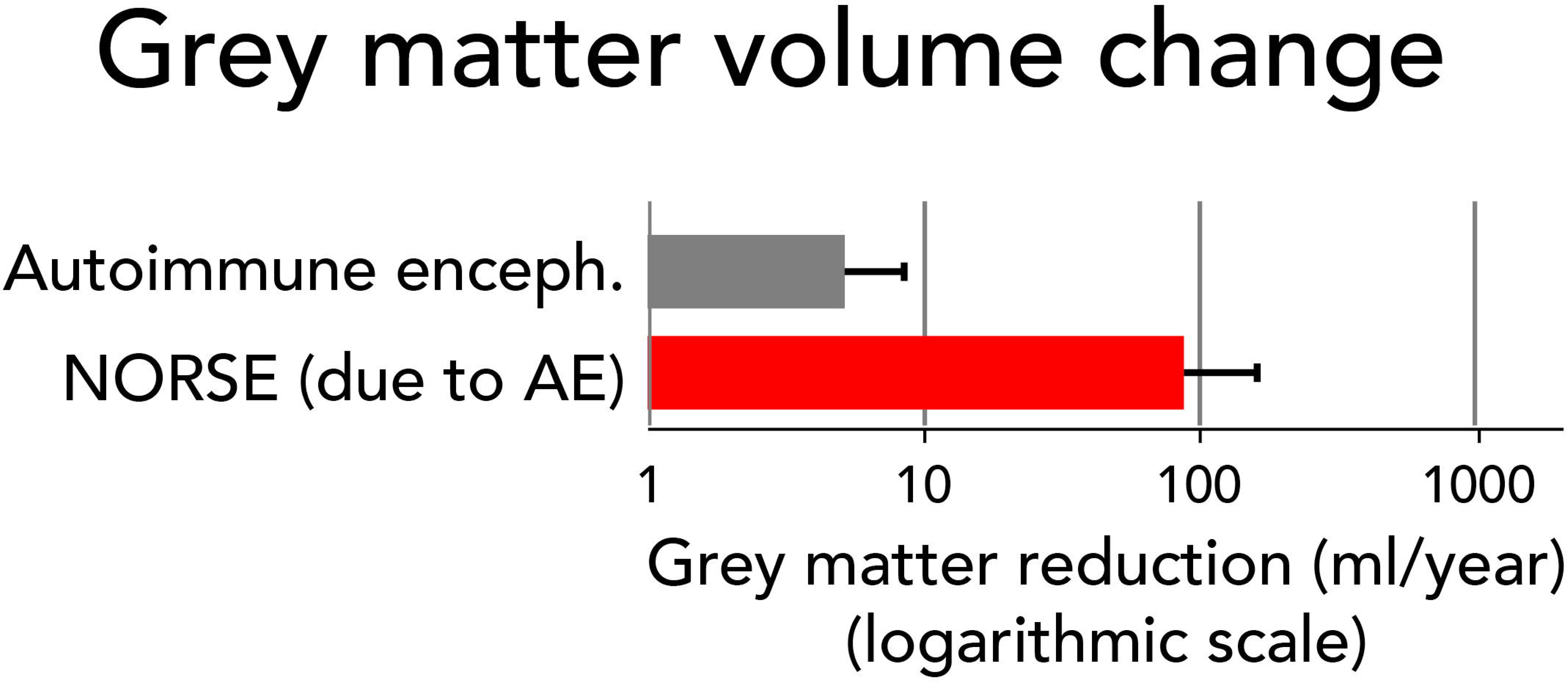
**Annualized morphometric changes in healthy aging, NORSE, and other neurological disorders.** Brain-wide plots depict annualized changes (log scale) in cortical thickness and subcortical volumes across groups: healthy controls **(A)**, individuals with epilepsy and brief seizures **(B)**, active autoimmune encephalitis **(C)**, mild cognitive impairment **(D)**, Alzheimer’s disease **(E)**, acute subarachnoid haemorrhage **(F)**, and NORSE **(G– I)**. NORSE data are shown for the full combined cohort **(G)**, discovery cohort **(H)**, and replication cohort **(I)**. The plots suggest a markedly accelerated brain shrinkage during NORSE compared to healthy aging and other neurological conditions, with consistent findings across both discovery and replication cohorts.

**Figure 2:**
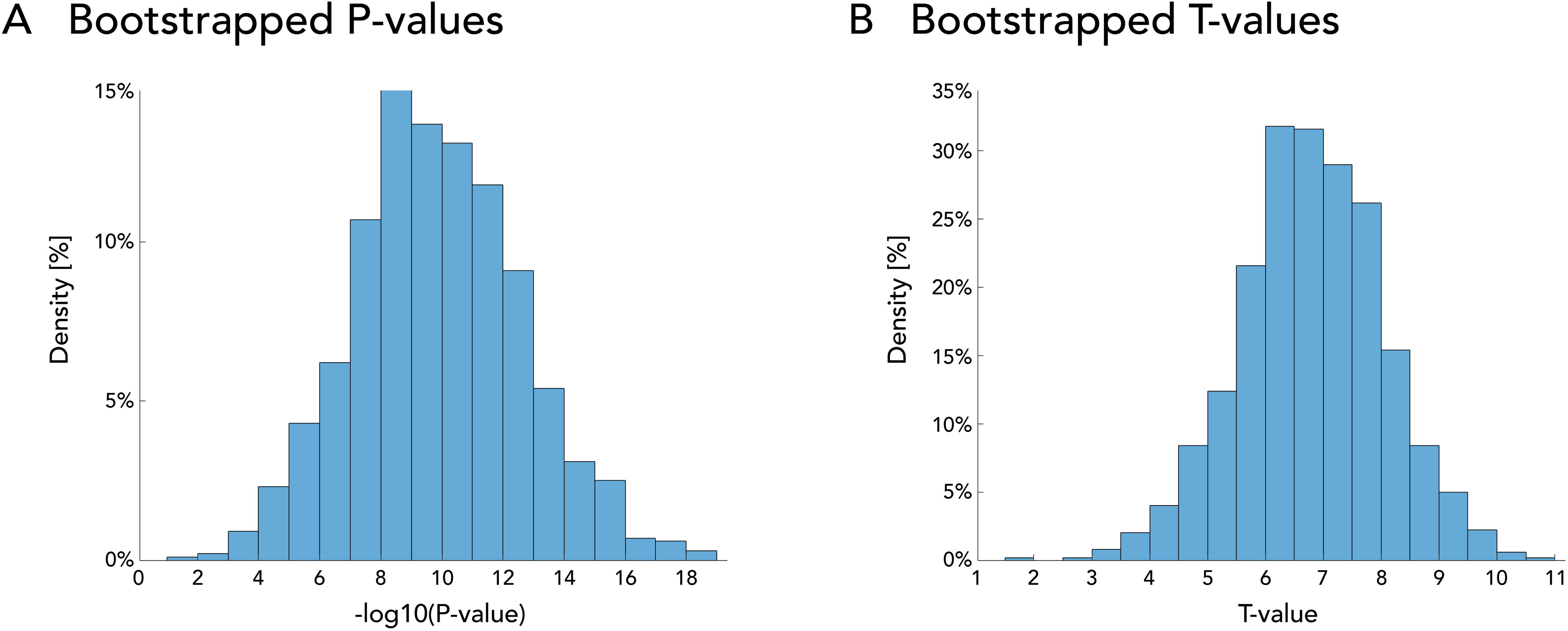
Association of NORSE with accelerated grey matter volume reduction. **(A, left)** Annualized grey matter volume (GMV) loss (ml/year, log scale) estimated using a linear mixed-effects model. Horizontal lines represent 95% confidence intervals (upper bounds shown). Asterisks indicate significantly faster GMV reduction compared to healthy aging. GMV loss during NORSE was approximately 80-fold higher than in healthy controls and 20-fold higher than in Alzheimer’s disease, with consistent findings across both the discovery and replication cohorts. **(A, right)** GMV plotted against age for the NORSE discovery (blue), replication (orange), and healthy control (grey) cohorts. Each dot represents a single MRI scan, with repeated scans from the same individual connected by lines. NORSE cases show rapid and substantial GMV decline, in contrast to the typically flat trajectory in healthy aging. **(B)** Brain maps showing regions of significantly accelerated cortical thinning and subcortical volume loss in NORSE compared to healthy controls. Blue clusters indicate atrophy; red clusters indicate thickness/volume increase. Results are shown for the full combined NORSE cohort (left) and separately for the discovery and replication cohorts (right). The findings highlight the widespread extent of NORSE-related brain atrophy. *NORSE, new onset refractory status epilepticus; T, thalamus; H, hippocampus; A, amygdala; Pa, pallidum; Pu, putamen; FWE, family-wise error*.

To demonstrate the generalisability of these findings, we observed similar GMV reduction rates (**Figure 2A**) in the multi-centre NORSE replication cohort (152.7 ± 78.4 ml/year, p=0.07 compared to subarachnoid haemorrhage; p<0.05 compared to all other groups) and in the combined full NORSE cohort (232.2 ± 46.1 ml/year, p<0.001 compared to all groups). To compare consistent aetiologies, NORSE due to autoimmune encephalitis had an 18-fold higher rate of GMV loss (88.3 ± 71.4 ml/year) compared to autoimmune encephalitis without SE (**Extended Data** Figure 1). Re-analysis using 1,000 bootstrapped random subsamples of the full NORSE cohort confirmed the stability of the statistical results (bootstrapped p-value 1.7 × 10r¹L, 95% CI 2.7 × 10⁻¹L – 2.8 × 10_⁻_L; bootstrapped t-value 6.8, 95% CI 4.2 – 9.2; **Extended Data** Figure 2). Results remained consistent across secondary analyses, including leave-one-cohort-out cross-validation, restricting to re-scans performed on the same scanner, and scans conducted on 3 Tesla or Siemens devices (**Extended Data Table 3**).

### Grey matter volume reduction in NORSE is irreversible

To assess whether brain atrophy in NORSE is reversible, we analysed data from six survivors who underwent MRI scans after the resolution of SE. Comparing mean GMV from the first (571L±L60Lml) to the last (559L±L41Lml) post-SE MRI, there was no evidence of recovery, i.e. no increase in GMV over time (t=-1.4; p=0.89; **Figure 3A**). The rate of GMV loss following NORSE (6.3L±L5.9Lml/year) was not significantly different from that seen in normal aging (p=0.24; **Figure 3B**).

**Figure 3:**
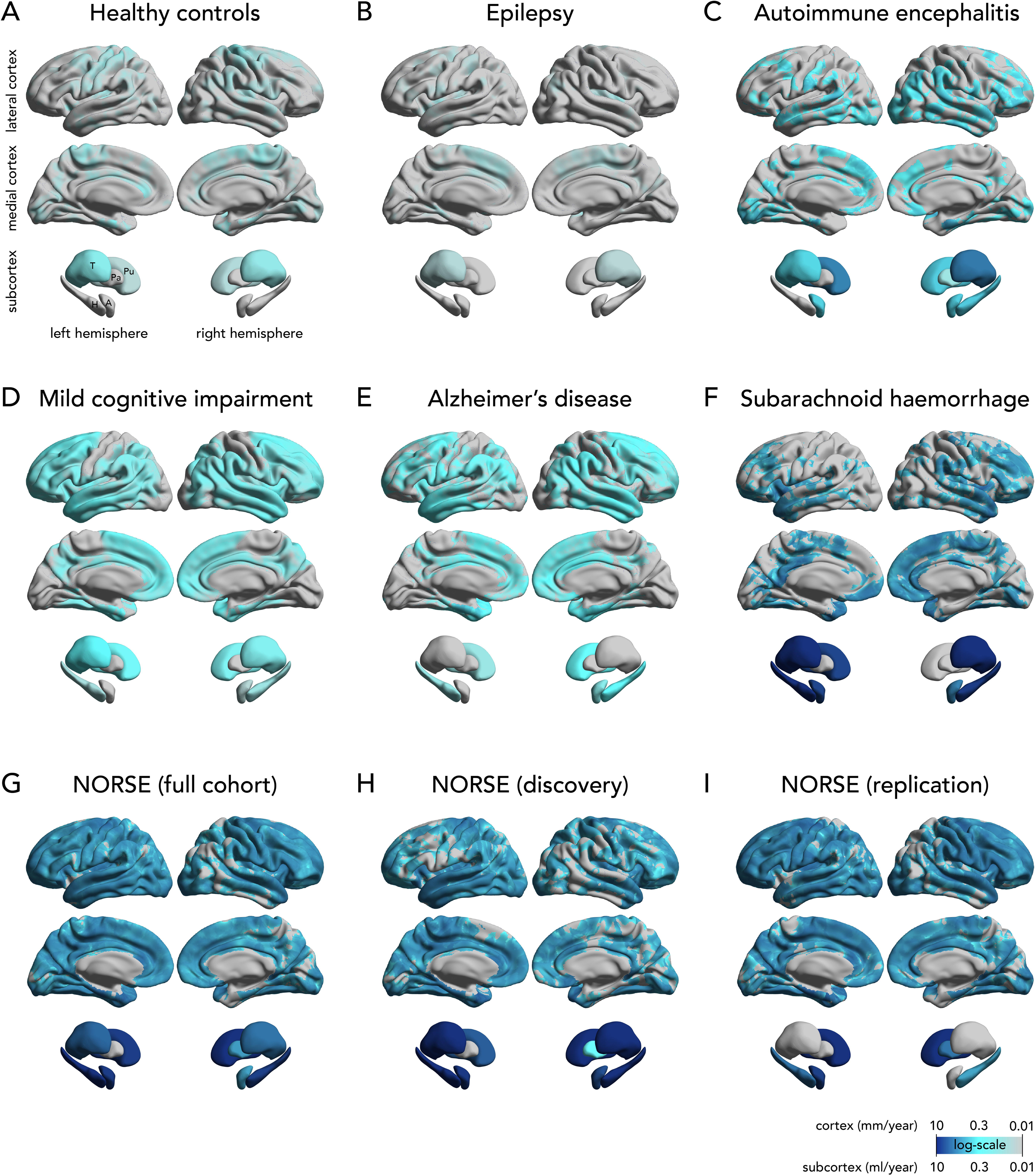
Reversibility of grey matter volume loss in NORSE. **(A)** Comparison of grey matter volume (GMV) between the first and last MRI scans obtained after resolution of NORSE. No increase in GMV was observed at follow-up, indicating a lack of reversal of atrophy. **(B)** Annualized GMV loss (ml/year, log scale) estimated using a linear mixed-effects model. Horizontal lines indicate 95% confidence intervals (upper bounds shown). While GMV loss was rapid during NORSE (p<0.001), the post-NORSE decline (p=0.24) did not differ from that observed in healthy controls, with no evidence of GMV recovery after SE cessation. **(C)** Relative grey matter volume (GMV) changes over time, referenced to the first available MRI scan. The x-axis is centred at the end of status epilepticus (end of SE = 0). Data are shown for six individuals with at least two MRI scans following SE resolution; each panel represents one subject. One subject (#3) had only post-SE scans. Each square denotes a single MRI scan, with longitudinal data points connected by lines. Scans obtained during NORSE are shown in red; post-NORSE scans in blue; transitions are indicated by orange lines. The data show GMV loss during NORSE, followed by a mostly stable trajectory after SE cessation, suggesting that the observed volume loss is largely irreversible.

Individual GMV trajectories (**Figure 3C**) showed a rapid decline during the NORSE episode *(red)*, followed by a largely stable course after SE resolution *(blue)*, with no visual evidence of reversal to baseline volumes.

### Rate of grey matter volume reduction correlates with clinical characteristics of NORSE

We analysed the association between clinical variables and the rate of brain shrinkage in the full NORSE cohort, adjusting for relevant covariables (**Figure 4**). The rate of GMV reduction was higher in cryptogenic NORSE compared to NORSE with a known aetiology (global: cryptogenic 345 ± 139 ml/year vs. known aetiology 124 ± 239 ml/year, p=0.03; regional: **Figure 4B**). Conversely, the rate was lower in NORSE caused by autoimmune encephalitis (global: autoimmune encephalitis 80 ± 234 ml/year vs. other aetiologies 325 ± 122 ml/year, p=0.02; regional: **Figure 4C**).

**Figure 4:**
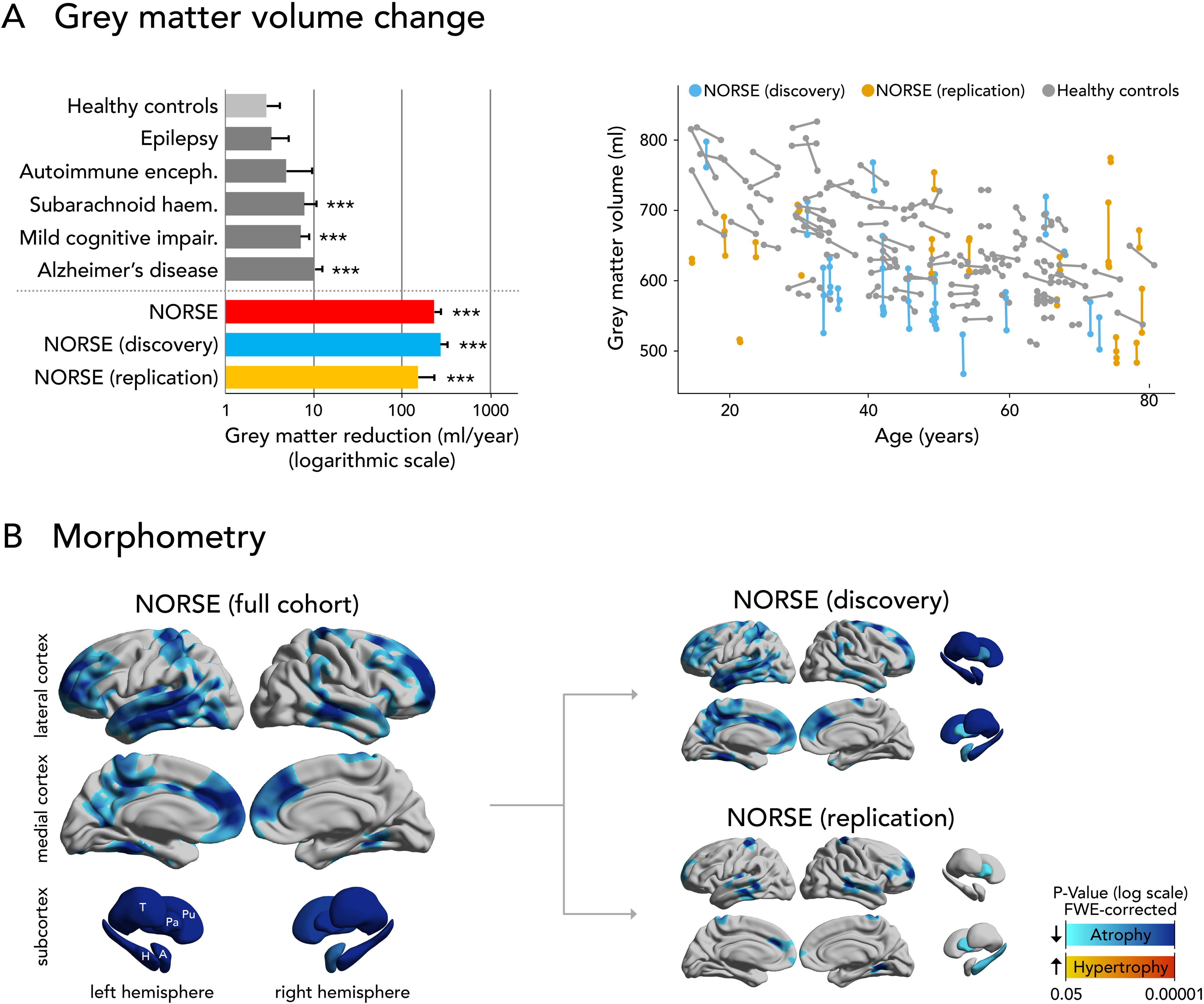
Association of grey matter volume loss during NORSE with clinical variables. **(A)** Annualized global grey matter volume (GMV) loss (ml/year, log scale) estimated using linear mixed-effects models. Horizontal lines indicate 95% confidence intervals (upper bounds shown). Asterisks denote statistically significant group differences. To visualize GMV loss across clinical subgroups, continuous variables (e.g., duration of status epilepticus) were dichotomized at the median for display purposes only (see Extended Data Table 5 for cut-offs); all statistical tests were based on continuous values. GMV loss was significantly greater in cryptogenic NORSE and significantly lower in cases due to autoimmune encephalitis. **(B–F)** Brain maps showing regional GMV loss associated with clinical characteristics. Blue clusters indicate atrophy; red clusters indicate relative thickness/volume increase. Results are shown for: **(B)** cryptogenic vs. non-cryptogenic aetiology; **(C)** autoimmune encephalitis vs. other aetiologies; **(D)** longer SE duration; **(E)** greater disability at last follow-up; and **(F)** death at last follow-up. *NORSE, new onset refractory status epilepticus; T, thalamus; H, hippocampus; A, amygdala; Pa, pallidum; Pu, putamen; FWE, family-wise error; FU, follow-up*.

A longer duration of NORSE was associated with regionally accelerated brain shrinkage (**Figure 4D**), although the global rate did not differ significantly (duration >19 days: 245 ± 249 ml/year; duration ≤19 days: 218 ± 146 ml/year; p=0.17). Higher disability (quantified with the modified Rankin Scale) at last follow-up and death were linked to accelerated volume reduction in the thalami (**Figure 4E-F**), while global rates remained unchanged. No other clinical variables showed significant associations with the rate of GMV reduction (**Extended Data Table 5**).

### Fluid biomarkers of neurodegeneration are markedly elevated during NORSE

Marked GMV decline may be mirrored by biological indicators of neurodegeneration. To complement the neuroimaging analyses, we measured fluid biomarkers of neuronal injury (cerebrospinal fluid Total Tau), axonal damage (serum neurofilament light chain, NfL), and astrocytic activation (serum glial fibrillary acidic protein, GFAP), adjusting all analyses for age, body mass index, and sex. All three biomarkers were elevated during NORSE (**Figure 5, Extended Data Table 6**), compared to two epilepsy cohorts with brief seizures (internal cohort: p values ranging from 0.004 to <0.001; external cohort for NfL and GFAP only: all p<0.001) and to patients with autoimmune encephalitis (all p<0.001). These findings remained significant after adjusting for potential confounders, such as non-specific systemic inflammation (serum C-reactive protein), renal function, and plasma exchange treatment.^45^

**Figure 5:**
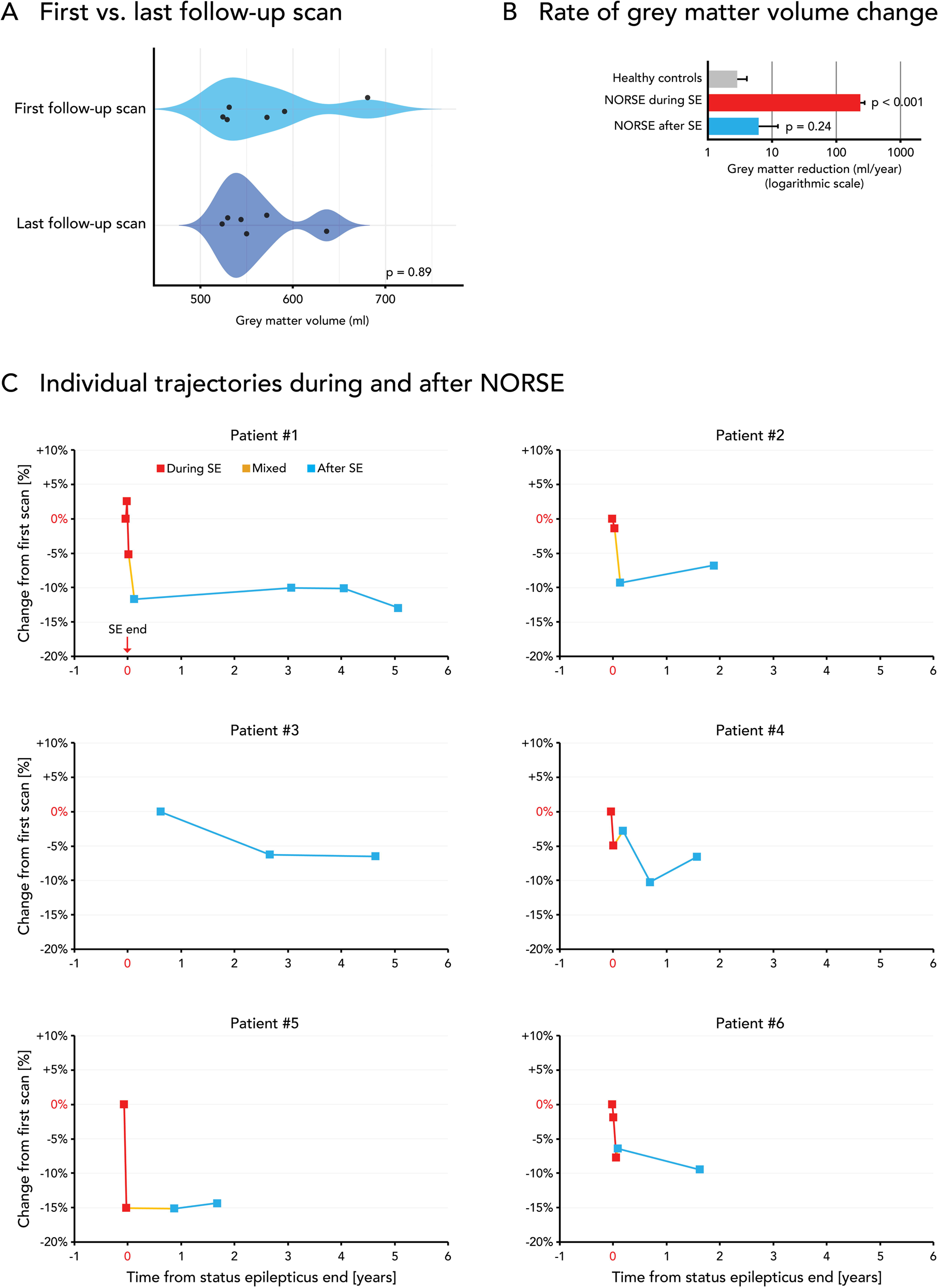
Fluid biomarkers of neurodegeneration are elevated during NORSE. **(A–C)** Levels of serum neurofilament light chain (NfL, **A**), cerebrospinal fluid (CSF) total Tau **(B)**, and serum glial fibrillary acidic protein (GFAP, **C**) in patients during NORSE, after NORSE, with autoimmune encephalitis, and in internal and external epilepsy cohorts with brief seizures. Horizontal lines represent 95% confidence intervals (upper bounds shown). Asterisks indicate statistically significant differences between measurements during NORSE and pairwise comparisons with all other groups. CSF total Tau was not available for the external epilepsy cohort. **(D–F)** Associations between fluid biomarkers during NORSE and clinical variables: **(D)** Higher NfL levels were associated with worse long-term functional outcome (modified Rankin Scale); **(E)** Total Tau was highest when measured early in the SE course; **(F)** GFAP correlated with greater SE severity. **(G–I)** Associations between fluid biomarker levels during NORSE and the rate of brain atrophy. Brain maps display regions where higher biomarker levels were associated with greater GMV loss (blue = atrophy; red = thickness/volume increase). Below each map, GMV loss estimates are shown using linear mixed-effects models. Continuous biomarker levels were dichotomized at the median for visualization only; all statistical analyses were based on continuous data. *NORSE, new onset refractory status epilepticus; NfL, neurofilament light chain; GFAP, glial fibrillary acidic protein; mRS, modified Rankin Scale; SE, status epilepticus; T, thalamus; H, hippocampus; A, amygdala; Pa, pallidum; Pu, putamen; FWE, family-wise error*.

All but one NORSE case showed abnormally elevated NfL during SE, based on both age-adjusted cutoffs^46^ and z-scores >2. After SE resolution, levels of NfL (p < 0.001), Total Tau (p = 0.007), and GFAP (p = 0.002) declined significantly and no longer differed from values in the epilepsy and encephalitis control groups (**Figure 5, Extended Data Table 6**). The single case with normal NfL levels during SE had an excellent functional outcome, returned fully to work, and reported no cognitive or neurological deficits following neurorehabilitation.

Higher levels of NfL (t = 1.4, p = 0.009; **Figure 5G**), Total Tau (t = 4.0, p < 0.001; **Figure 5H**), and GFAP (t = 3.0, p = 0.004; **Figure 5I**) were associated with more rapid GMV decline on serial MRI (**Extended Data Table 7**).

Elevated NfL during SE was also associated with worse long-term functional outcome (modified Rankin Scale; t = 3.3, p = 0.02; **Figure 5D**). Total Tau levels were highest when measured early in the SE course (t = –4.2, p = 0.004; **Figure 5E**), consistent with its rapid-release kinetics, whereas no such temporal association was observed for NfL or GFAP. GFAP levels correlated with greater SE severity (t = 5.3, p = 0.001; **Figure 5F**) and fewer anaesthetic drugs administered (t = –2.5, p = 0.04).

### Histological specimen show neuronal loss and gliosis

Two subjects from the discovery cohort underwent cortical biopsy, which was preserved for histological analysis. In both cases, immunohistochemistry using antibodies against neuronal nuclear antigen (NeuN) and the astrocytic marker GFAP revealed severe neuronal loss and reactive astrogliosis, respectively (**Figures 6E and 6J**). Full case descriptions are provided in the **Supplementary Information**.

**Figure 6:**
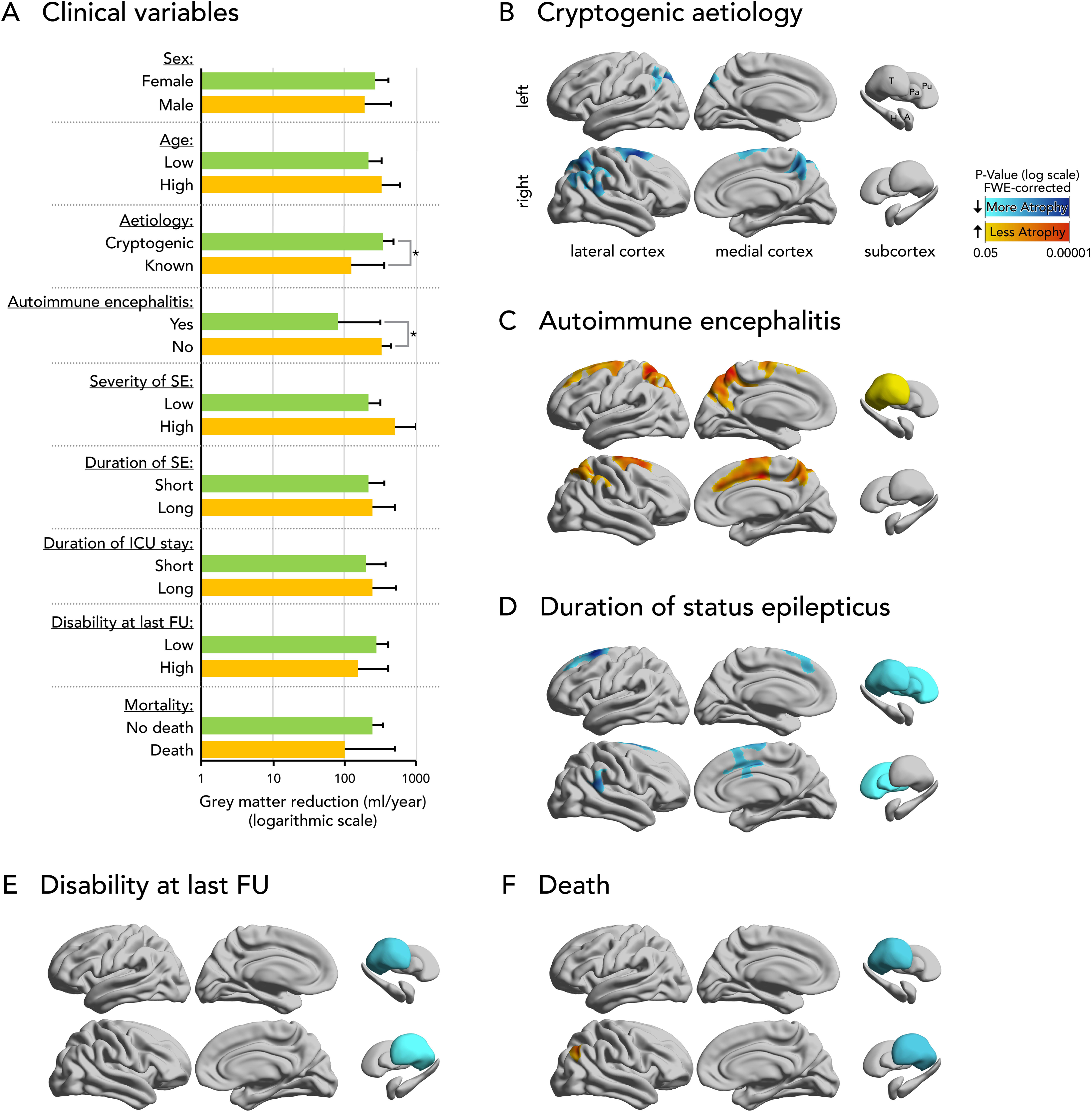
Two representative NORSE cases with serial MRI, fluid biomarkers, and neuropathology data. **(A, F)** Selected axial slices from the first, third, and fifth MRI scans performed during NORSE in each case. Progressive enlargement of cerebrospinal fluid spaces, consistent with brain atrophy, is evident in the final scan compared to the initial one. **(B, G)** Relative changes in brain morphometry over time from SE onset, referenced to the first available MRI scan. Each square represents one MRI, with scans from the same individual connected by lines. Both cases showed rapid grey matter volume loss, particularly affecting subcortical structures, with minimal change in white matter volume. **(C, H)** Annualized whole-brain morphometric changes between the first and last MRI during NORSE, mapped onto cortical and subcortical templates. Brain shrinkage was bilateral and widespread in both cases. **(D, I)** Longitudinal trajectories of fluid biomarkers of neurodegeneration (neurofilament light chain, NfL; glial fibrillary acidic protein, GFAP; Total Tau), referenced to the internal epilepsy cohort mean (log-scale, k-fold increase from epilepsy mean). **(E, J)** Neuropathological findings compared with a representative healthy cortex (bottom panel). In Case 1 **(E)**, cortical tissues showed decreased numbers of neuronal nuclear antigen (NeuN) positive neurons with preserved layering with Luxol fast blue staining and marked GFAP-positive reactive astrogliosis. In Case 2 **(F)** there were several fragments of cerebral cortex and subcortical white matter with reduced neuronal numbers and reactive astrogliosis. *NORSE, new onset refractory status epilepticus; NfL, neurofilament light chain; GFAP, glial fibrillary acidic protein; CSF, cerebrospinal fluid;* neuronal nuclear antigen, NeuN; *T, thalamus; H, hippocampus; A, amygdala; Pa, pallidum; Pu, putamen*.

Case 1 (**Figures 6A-E**) was a previously healthy male in his early forties who developed cryptogenic NORSE lasting 3.3 months following a brief febrile illness. He was discharged to rehabilitation in an awake but severely dependent state; however, he experienced a relapse of SE and subsequently died.

Case 2 (**Figures 6F-J**) was a previously healthy female in her late forties who experienced cryptogenic NORSE lasting 3 months, also following a brief febrile illness. Several years later, she continues to suffer from pharmacoresistant focal epilepsy and severe cognitive deficits. While she can communicate and walk unaided for a few steps, her quality of life remains significantly impacted.

Brain morphometry demonstrated substantial GMV reductions of 17.1% and 9.4% between the first and last scans in Case 1 and Case 2, respectively (**Figures 6B and 6G**). GMV reduction appeared to accelerate most rapidly during the initial 0.5 to 1 month of NORSE and stabilized thereafter. These changes predominantly affected subcortical GMV (Case 1, −23.4%; Case 2, −28.1%) more than cortical thickness (Case 1, −8.1%; Case 2, −4.5%) and had minimal impact on white matter volume.

The atrophy was distributed across widespread bi-hemispheric areas, encompassing almost the entire cerebral cortex and subcortex (**Figures 6C and 6H**).

Fluid biomarkers of neurodegeneration were assessed in Case 1 at four time points (**Figure 6D**). NfL levels were consistently elevated, showing a 9-to 16-fold increase compared to the internal epilepsy cohort. Total Tau was markedly elevated in the early phase of SE (74-fold increase) but declined over time (2.7-fold at the final sample). GFAP levels were not elevated. In Case 2, a single serum measurement one week after NORSE onset, showed a 16-fold increase in NfL and a 4-fold increase in GFAP (**Figure 6I**).

These cases point towards correlation of GMV loss, cortical neuronal damage, and concurrent glial reactive changes with consistent changes in CSF and blood markers during NORSE.

## Discussion

This study shows that the rate of brain shrinkage during NORSE is markedly greater than in normal aging (80-fold), epilepsy with brief seizures (70-fold), and common neurodegenerative conditions (20–30-fold). It also exceeds rates observed in inflammatory conditions such as autoimmune encephalitis (50-fold) and in scenarios requiring prolonged ICU treatment, such as subarachnoid haemorrhage (30-fold), indicating a direct effect of SE on brain structure. These findings were consistent across multicentre external validation, multiple methodologies (brain MRI, fluid biomarkers, and histopathology), and several secondary analyses. SE-related atrophy was widespread, irreversible, and associated with SE duration and long-term outcomes. The marked elevation of fluid biomarkers of neurodegeneration during NORSE, which normalized following SE resolution, suggests cellular damage consistent with the observed MRI findings. Collectively, these results underscore NORSE as a neurological emergency associated with irreversible brain injury and provide a framework for evaluating neuroprotective interventions in humans.

The concept that SE can cause brain damage is key to our understanding of the disorder. Its current duration-based definition^13^ incorporates evidence from experimental models indicating that prolonged seizures lead to neuronal injury,^5,14,15^ whereas human evidence has been comparatively scarce. Our findings address this gap by showing markedly accelerated brain shrinkage in NORSE. Because NORSE often arises without significant pre-existing neurological disorders, it reduces the confounders typically present in other SE subtypes and enables a clearer assessment of seizure-related damage. However, potential contributions from an underlying immune-mediated aetiology, prolonged ICU stay, and exposure to neurotoxic treatments^47^ must also be considered when interpreting these findings, as discussed below.

MRI-based measures of GMV and cortical thickness capture macroscopic correlates of microscopic processes such as neuronal loss, synaptic and dendritic pruning, and glial alterations.^48^ While these measures do not directly quantify neuronal loss, their decline is a well-established proxy for neurodegeneration and is observed across multiple neurodegenerative disorders.^49^ In our study, the rate of GMV loss correlated with fluid biomarkers of neurodegeneration (**Figures 5G, 5H and 5I**), reinforcing the hypothesis that MRI-detected atrophy reflects an underlying neurodegenerative process. Similarly, a previous study found that elevated CSF Tau was associated with hippocampal atrophy after NORSE.^34^ Histological analyses in two participants further confirmed neuronal loss and reactive astrogliosis during NORSE (**Figures 6E and 6J**), and a systematic review reported neuronal loss in 11 of 16 NORSE autopsy cases.^50^ Notably, brain atrophy did not rebound after the acute phase (**Figure 3**), suggesting that the structural damage is both substantial and irreversible. The magnitude of GMV decline observed in our study aligns with previous reports showing a 16-23% decrease in brain-ventricle ratio^29,31^ and 23% hippocampal atrophy^30^ in individuals with peri-ictal MRI abnormalities, nonconvulsive seizures, and super-refractory SE.

Alternative explanations, such as progressive hydrocephalus, transient inflammatory or toxic changes, or resolution of initial brain swelling, are unlikely to fully account for the timing, magnitude, spatial distribution, irreversibility, and grey matter selectivity of the observed changes. Furthermore, baseline MRI measurements were within normal ranges for age (**Figure 2B**), showing no signs of initial swelling, and clinical signs of hydrocephalus were not reported in most NORSE cases.

Several mechanisms may contribute to neurodegeneration in NORSE. Firstly, SE may directly induce excitotoxic injury through excessive glutamate release and NMDA receptor activation.^18,19,51^ Secondly, NORSE is believed to involve immune-mediated mechanisms that can promote neurodegeneration.^52^ However, atrophy rates and neurodegeneration biomarker levels were markedly higher in NORSE than in autoimmune encephalitis. Even among cases of NORSE in association with autoimmune encephalitis, the rate of GMV loss was 18-fold higher compared to autoimmune encephalitis without SE (**Extended Data** Figure 1), supporting the hypothesis of a direct contribution of SE to brain atrophy, independent of underlying aetiology. Thirdly, NORSE often requires prolonged sedation and treatment with potentially neurotoxic medications.^47^ Yet, atrophy rates during NORSE were 30-fold higher than in subarachnoid haemorrhage, a condition similarly linked to extended ICU stays and sedation. Together, these findings indicate that neither the underlying aetiology of NORSE nor ICU-related factors fully explain the degree of brain shrinkage. Instead, it is possible that a substantial proportion of neurodegeneration in NORSE results from the direct effects of prolonged SE.

Whether the rate of neurodegeneration predicts long-term outcomes in NORSE remains uncertain, however, our findings suggest potential associations. Accelerated thalamic atrophy was linked to worse functional outcomes, possibly reflecting more severe excitotoxic injury in this key hub for arousal and motor-cognitive integration.^53^ Neuropathology-confirmed bilateral thalamic neurodegeneration has been previously described following SE. Similarly, higher NfL levels correlated with poor functional outcomes, underscoring the relevance of axonal damage.^54^ Future studies with larger cohorts should investigate the relationship between brain atrophy and cognitive or functional outcomes, although establishing a pre-SE cognitive baseline remains challenging, as patients rarely undergo assessment prior to NORSE onset.

Additionally, patients with cryptogenic NORSE exhibited a faster rate of brain shrinkage. This may reflect the lack of a known targetable cause. In other words, when the aetiology is known, treatment can be tailored more precisely and may better protect against brain damage.

This study has several limitations. Firstly, the retrospective design limits the inference of causality. Because serial MRIs in NORSE were performed for clinical diagnostics, our analyses may be biased toward more severe, prolonged, and cryptogenic cases. However, the included cohort was representative of consecutive NORSE cases at our primary site (**Extended Data Table 3**), suggesting only minimal bias. Secondly, some NORSE patients underwent MRI scans on different scanners, potentially introducing variability. Yet, secondary analyses restricted to the same scanning equipment yielded consistent results (**Extended Data Table 4**), making scanner-related bias unlikely. Thirdly, the epileptic focus was unknown in most NORSE cases, preventing us from correlating localized brain damage with proximity or connectivity to the focus. Moreover, prolonged SE may involve large-scale networks rather than a single focus, which could explain the extensive structural brain changes seen during NORSE. Fourthly, NfL and GFAP measurements in the external epilepsy cohort were obtained using a different assay, which may limit direct comparability. However, fluid biomarker levels during NORSE remained significantly elevated when compared to the internal epilepsy and autoimmune encephalitis cohorts, which were analysed using the same assay. Fifthly, although our sample size is relatively large in the context of NORSE research, larger cohorts with standardized cognitive assessments are needed to clarify how medications influence both brain atrophy and cognitive outcomes. Finally, we focused exclusively on NORSE, and it remains unclear whether our findings generalize to other forms of SE. However, evidence from other studies suggests that brain damage may also occur in other forms of refractory SE.^29–33,35,36^

In conclusion, we observed markedly accelerated and irreversible brain atrophy during NORSE, likely driven by the toxic effects of prolonged seizures, underlying aetiologies, and administered treatments. These results underscore the importance of rapid diagnosis and early termination of NORSE to minimize brain damage. Serial MRI and fluid biomarkers offer a promising framework for investigating neurodegeneration in SE and may represent a monitoring tool to guide and direct neuroprotective interventions.

## Online Methods

### Participants and datasets

Participants and datasets are described briefly here, with additional details in the **Supplementary Information**. Power analysis (α = 0.05, two-sided; 80% power) indicated that 14 participants per group were sufficient to detect an expected 15-percentage-point difference in rate of GMV decline^29–31^ between cases and controls using a linear mixed-effects model assuming two repeated measurements per subject (see **Supplementary Information** for details).

### NORSE cohort

NORSE was defined according to the current consensus definition.^37^ For the discovery cohort, we screened adults with SE admitted to the Neurocritical Care Unit of the University Hospital Zurich, Switzerland, between January 2010 and December 2023. We included individuals who had at least two MRI scans meeting our quality criteria (see below) separated by at least one week, with at least one scan obtained during NORSE. Scan pairs overlapping with the duration of NORSE were used for the main analyses, whereas scan pairs obtained after NORSE were used to assess reversibility (**Figure 3**).

For the replication cohort, we screened adults from six external sites—University Hospital of Basel (Switzerland), Hôpitaux Universitaires de Genève (Switzerland), University College London (United Kingdom), LMU University Hospital Munich (Germany), Christian Doppler University Hospital (Salzburg, Austria), and Kantonsspital St. Gallen (Switzerland) using the same inclusion criteria as in the discovery cohort. The descriptions of each cohort are provided in the **Supplementary Information**.

Severity of SE was measured using the status epilepticus severity score (STESS).^55^ Mortality and disability, measured using the modified Rankin scale, were determined at last available follow-up.

### Comparison cohorts

We also included subjects with serial MRI to compare morphometric changes in subjects with NORSE versus normal aging (healthy controls), brief epileptic seizures without SE, and other neurological conditions associated with acute and chronic neurodegeneration, such as autoimmune encephalitis, subarachnoid haemorrhage, mild cognitive impairment, and Alzheimer’s disease. This allowed us to estimate the partial effects of underlying inflammatory processes, prolonged ICU treatment and underlying normal and pathological aging on brain atrophy and to determine whether the rate and degree of morphometrical changes during NORSE differs from that of these conditions.

Participants in all comparison cohorts had no current or prior status epilepticus and contributed ≥ two MRI scans that satisfied our predefined quality criteria, acquired at least one week apart. A concise description of each cohort follows; comprehensive details are provided in the **Supplementary Information** and **Extended Data Table 2**.

### Epilepsy with brief seizures

We included adults with established focal or generalized epilepsy, as defined by the International League Against Epilepsy (ILAE) practical clinical definition,^56^ who had no history of SE and underwent serial MRI 3.5 years apart. Participants were prospectively recruited between June 1995 and May 1997 as part of a prospective population-based study in Buckinghamshire, United Kingdom with 207,553 residents.^57^ Brief seizures have been defined as seizures that do not meet the definition of SE.^13^

### Autoimmune encephalitis

We included patients with definite autoimmune encephalitis according to current criteria^58^ treated at the University Hospital Zurich, Switzerland, between January 2013 and December 2024 who had at least two MRI scans one week or more apart, covering at least part of the active disease phase and no SE. Patients were categorized as “active” if suffering from an acute disease, a relapse or a chronic progressive disease course.

### Subarachnoid haemorrhage

We included patients with neuroimaging-confirmed acute subarachnoid haemorrhage who required neurocritical care at the University Hospital Zurich, Switzerland, between January 2017 and August 2024, had no SE, and received serial MRI at least one week apart during admission. We considered the disease “severe” if it required monitoring on our neurocritical care unit (mean 24 ± 15 days ICU admission).

### Alzheimer’s disease and mild cognitive impairment

We analysed longitudinal T1-weighted MRI data of people with Alzheimer’s disease or mild cognitive impairment from the Alzheimer’s Disease Neuroimaging Initiative (ADNI) Phase 1 study. ADNI-1 was a multi-centre natural-history study that has prospectively enrolled ∼800 adults aged 55–90 years at ∼50 North-American sites between 2005 and 2007. Participants underwent certified high-resolution 3D T1 imaging on harmonised MRI scanners every 6–12 months (Alzheimer’s disease: baseline, 6, 12, 24 months; mild cognitive impairment: baseline, 6, 12, 24, 36 months). Diagnostic-group specific criteria required a Mini-Mental State Examination of 24–30 for MCI, 20–26 for AD, and a Clinical Dementia Rating of 0.5 for MCI and 0.5–1.0 for AD. We included individuals who contributed ≥ 2 scans that met ADNI quality-control criteria.

The ADNI was launched in 2003 as a public-private partnership, led by Principal Investigator Michael W. Weiner, MD. The primary goal of ADNI has been to test whether MRI, positron emission tomography, other biological markers, and clinical and neuropsychological assessment can be combined to measure the progression of mild cognitive impairment and early Alzheimer’s disease.

### Healthy controls

Healthy controls were obtained from three sources: (1) a population-based study in Buckinghamshire, United Kingdom (the same study as the epilepsy cohort),^57^ (2) the Neuromorphometry by Computer Algorithm Chicago (NMorphCH) study,^59^ and (3) the Parkinson Progression Marker Initiative (PPMI) study.^60^ These controls were propensity-score matched with the full NORSE cohort for age and sex at a 3:1 ratio (see **Supplementary Information** for details). Throughout the manuscript, “normal aging” refers to the brain-morphometry change rates observed in the healthy control group.

### MRI data

Minimal quality criteria for eligible MRI scans included a three-dimensional T1-weighted sequence with high grey–white matter contrast (e.g., Magnetization Prepared Rapid Gradient Echo, Spoiled Gradient Echo, or Turbo Field Echo), and an isotropic or near-isotropic voxel size of 1.5 mm or smaller. We excluded MRIs with excessive motion or other artifacts, space occupying lesions likely to disrupt MRI preprocessing, or posttraumatic/postoperative cavities and subjects with clinical diagnosis of other neurodegenerative conditions aside from the above. All raw scans were visually inspected by MG for quality control. We excluded any scans that did not meet the defined quality standards or were otherwise unusable (e.g., due to motion artifacts).

### MRI preprocessing

We estimated total intracranial volume, grey and white matter volumes, cortical thickness, and regional subcortical volumes for all scans in all cohorts using the fully automated,^61^ validated,^61–63^ and reliable^62,63^ Computational Anatomy Toolbox (CAT12; Jena University Hospital, Germany; https://neuro-jena.github.io/cat/) within Statistical Parametric Mapping 12 (SPM12; Wellcome Centre for Human Neuroimaging, United Kingdom ;https://www.fil.ion.ucl.ac.uk/spm/software/spm12/) running in MATLAB R2023b (The MathWorks, Natick, MA, USA) with an inverse-consistent longitudinal registration approach. The CAT12 toolbox showed excellent test-retest reliability (R^2^ = 0.986) and was validated against other cortical surface reconstruction methods, showing fewer thickness measurement errors than comparable approaches.^62,63^

Cortical thickness was computed using CAT12’s projection-based thickness method using standard settings, which derives white matter distance from tissue segmentation obtained using geodesic shooting and projects local maxima of thickness to grey matter voxels, effectively accounting for partial volume effects and sulcal geometry. The pipeline includes topology correction using spherical harmonics, spherical mapping to a common coordinate system, diffeomorphic surface registration for inter-subject alignment, and subsequent smoothing with a 12 mm surface-based kernel.

Subcortical regional volumes in five regions (hippocampus, amygdala, thalamus, pallidum, and putamen) were calculated by projecting grey matter volume onto the Hammers brain atlas in template space.^64^

All data underwent CAT12-based quality control, and scans with misalignment, misregistration, or inaccurate estimations were excluded. Image quality was assessed by evaluating image noise, inhomogeneities, and resolution within the CAT12 quality assurance framework.

### Fluid biomarkers

We reviewed the institutional cerebrospinal fluid (CSF) and serum biobank at the University Hospital Zurich for samples from individuals with NORSE and patients with autoimmune encephalitis that were included in the MRI study. We also included a consecutive cohort of patients with non-lesional epilepsy who underwent CSF testing as part of their diagnostic work-up between 2022 and 2024. All samples were stored in polypropylene tubes at −80 °C. Prior to analysis, samples were thawed at room temperature and homogenized using low speed vortexing for at least 30 seconds.

Serum concentrations of neurofilament light chain (NfL) and glial fibrillary acidic protein (GFAP) were measured using the Single Molecule Array (Simoa) Neurology 2-Plex B assay on the SR-X Analyzer (Quanterix Corp., Billerica, MA, USA). CSF total Tau concentrations were quantified using the Lumipulse G Total Tau assay (Fujirebio Inc., Tokyo, Japan). All assays were performed according to the manufacturers’ protocols. In cases where Total Tau concentrations exceeded the upper limit of quantification, samples were serially diluted, re-pipetted, and vortexed to ensure complete homogenization before reanalysis.

As an additional comparison group, we included previously acquired plasma NfL and GFAP data from the Prospective Regional Epilepsy Database and Biobank for Individualized Clinical Treatment (PREDICT), a regional biobank study based in Västra Götaland, Sweden (clinicaltrials.gov, NCT04559919).^65^ Following recruitment, blood samples were centrifuged at 2000×g for 10 minutes at room temperature, aliquoted, and stored at −80 °C until analysis. NfL and GFAP levels in this cohort were measured using the Simoa N4PB kit on an HD-1 Analyzer (Quanterix Corp., Billerica, MA, USA).

### Histology and case reports

Two individuals with NORSE from the discovery cohort underwent brain biopsies. We briefly describe their cases in the main manuscript, with additional details provided in the **Supplementary Information**. We assessed the relative changes in grey and white matter volumes, mean cortical thickness, and subcortical volumes relative to the first available scan. We also calculated the annualised rate of vertex-wise cortical thickness and subcortical volume changes between the first and last scans during NORSE. Fluid biomarkers were reported as a fold-increase in comparison with the median of the internal epilepsy cohort that was analysed using the same assay.

### Neuropathology

Sections were performed during routine diagnostics using standard protocols. 2 µm thick FFPE and frozen sections were cut on a HP microtome (Leica Biosystems). H&E stains were performed using a Tissue-Tek® Prisma™ automated slide stainer (Sakura Finetek), Luxol-Fast-Blue/Nissl stain was performed by hand (all staining solutions from Artechemis). Immunohistochemistry was performed using OptiView DAB IHC Detection Kit (Roche) or Bond Detection Kit (Leica Biosystems) on a Ventana (Roche) or a BOND-III Leica Biosystems) fully automated immunohistochemistry staining system. The following primary antibodies were used: mouse-anti-human CD68 (clone PG-M1, Dako Omnis, 1:100 on BOND-III), rabbit-anti-human GFAP (polyclonal, Dako Omnis, 1:10000 on Ventana), mouse-anti-human NeuN (monoclonal, clone A60, Merck Millipore, 1:4000 on Bond). Slides were investigated and images acquired with a BX43 Manual System Olympus microscope and the cellSens entry software (Olympus, Tokio, Japan). Furthermore, overview images were acquired using a Hamamatsu C9600 digital slide scanner (Hamamatsu Photonics, Hamamatsu, Japan).

### Statistics

#### Main statistical analyses

We estimated the rate of GMV change using linear mixed-effects models, a flexible framework for longitudinal analysis of multiple repeated measurements per subject with irregular measurement intervals. To assess differences between groups (e.g., NORSE vs. controls) or the influence of clinical variables (e.g., SE duration) on morphometric parameters over time, we tested the interaction between the variable of interest (group allocation or clinical variable) and the interscan interval. All models included subjects as a random effect and fixed effects of age at first scan, interscan interval, sex, group, and total intracranial volume. This approach enabled us to capture within-subject changes over time while adjusting for baseline demographic differences and varying interscan intervals. We classified findings as group-level brain atrophy when statistical tests showed significant global or regional GMV loss or cortical thinning compared with age- and sex-matched healthy controls modelling normal aging.

To reduce confounding, we controlled analyses of clinical variables and medication effects for duration of SE, severity of status epilepticus, NORSE aetiology, number of antiseizure medications and sedatives, and site location. For continuous or ordinal variables, the rate of morphological change was displayed by splitting the data at the median (for illustration only), while statistical analyses were performed on the unsplit data.

Categorical demographic variables are presented as N (%) and were analysed using Fisher’s exact test. Continuous variables are presented as mean ± standard deviation (SD) and were analysed using the Mann–Whitney U-test. We performed statistical analyses using R 4.4.1 (R Foundation for Statistical Computing).

#### Cortical and subcortical morphometric analyses

We assessed the spatial extent of our findings by applying the same linear mixed-effects models to vertex-wise cortical thickness and regional subcortical volumes using SurfStat (https://www.math.mcgill.ca/keith/surfstat/) in MATLAB. Morphometric findings were reported at p < 0.05, corrected for multiple comparisons using random field theory for nonisotropic images at the cluster level^66^ for cortical thickness, and Bonferroni correction for subcortical volumes.

Plots of annualized morphometric changes (**Figure 1**) were calculated by taking the absolute difference in vertex-wise cortical thickness and subcortical volumes for each aligned scan pair and dividing by the interscan interval. Mean annualized changes were then displayed for each cohort on a representative brain template.

#### Secondary analyses

To test the robustness of our main findings, we repeated the GMV-based analyses in a leave-one-cohort-out cross-validation approach and in relevant subgroups. To assess the stability of our statistics and estimate uncertainty, we performed bootstrapping by re-estimating the model 1,000 times with randomly selected subsamples, then calculated the distribution of p and t values.

#### Reversibility of morphometric findings

To assess the reversibility of GMV reduction after NORSE, we included six individuals who had at least two MRI scans following SE cessation and calculated the differences in global GMV between the first and last scan after SE using a paired t test. We then applied linear mixed-effects models, as described above, to compare the rate and magnitude of GMV change after SE cessation with that of healthy controls. We also plotted the relative GMV change with respect to the first available scan, using colour coding to indicate whether the interscan intervals occurred during or after status epilepticus.

#### Fluid biomarker analyses

To generate interpretable NfL data adjusted for age, body mass index (BMI), and assay platform, raw NfL concentrations were converted to z scores using a previously validated method (https://shiny.dkfbasel.ch/baselnflreference/).^67^ As no equivalent z-scoring method is currently available for Total Tau and GFAP, raw values were used for these markers, with confounders accounted for in the statistical models.

Between-group differences in fluid biomarkers were assessed using linear mixed-effects models, adjusted for age, sex, and BMI, with subjects included as a random effect to account for repeated measures. For NfL z scores, adjustments for age and BMI were not applied, as these variables were already incorporated into the scoring algorithm. As secondary analyses, models were refitted with additional adjustments for non-specific systemic inflammation (C-reactive protein), renal function, and plasma exchange treatment because it has been reported that these factors may affect NfL values.^45^ Bonferroni correction was applied for multiple testing.

Associations between fluid biomarkers and clinical variables were evaluated using linear mixed-effects models adjusted for age, sex, BMI, SE duration, SE severity (STESS), number of antiseizure and anaesthetic medications, NORSE aetiology, and time from SE onset to sample collection. Associations between fluid biomarkers and morphometric measures were assessed using the same statistical modelling approach applied to other continuous variables, as described above.

### Data availability

Data are available upon reasonable request with formal applications submitted to the respective cohort to protect patient sensitive data. Data from specific cohorts can be requested from the following cohort leaders: University Hospital Zurich, MG; University Hospital of Basel, UF; Hôpitaux Universitaires de Genève, PdS; University College London, SR; LMU University Hospital, MLS; Christian Doppler University Hospital Salzburg, GK; Kantonsspital St. Gallen, DZ; Prospective Regional Epilepsy Database and Biobank for Individualized Clinical Treatment, JZ. The ADNI data can be requested at the ADNI database (https://adni.loni.usc.edu/).

Healthy control data from the Neuromorphometry by Computer Algorithm Chicago and Parkinson Progression Marker Initiative can be requested at https://schizconnect.org and https://www.ppmi-info.org respectively.

### Code availability

The analysis code is publicly available through GitHub (https://github.com/mgalovic/norse).

## Sources of funding

The current study was funded by the Swiss National Science Foundation (Grant number 215619) and the Betty & David Koetser Foundation for Brain Research and supported by the National Institute for Health and Care Research University College London Hospitals Biomedical Research Centre.

## Disclosures

Cyril Simmen received research grants from the Betty & David Koetser Foundation for Brain Research. Marian Galovic reports compensation from Angelini Pharma, Bial, Eisai, Neuraxpharm, and UCB for consultant services. Johan Zelano reports speaker honoraria from UCB, Eisai, Angelini Pharma, and Orion Pharma and as an employee of Sahlgrenska University Hospital being investigator in clinical trials sponsored by Bial, SK Life Science, UCB, and GW Pharma. Nils Briel received research grants from Fondation suisse de recherche sur les maladies musculaires and ETH Zurich MedLab Fellowship programme, none of which are related to this work. Anton Schmick received travel grants from Novo Nordisk and Merck Pharma, none of which is related to this work. Marina Herwerth was supported by the Swiss National Science Foundation (PZ00’3_216616/1), by the Olga-Mayenfisch-Foundation (2024) and by Betty and David Koetser Foundation. Patrick Roth has received honoraria for lectures or advisory board participation from Alexion, Bristol-Myers Squibb, Boehringer Ingelheim, CDR-Life, Debiopharm, Galapagos, Laminar, Midatech Pharma, Novartis, Novocure, OM Pharma, QED, Roche, Sanofi and Servier and research support from Merck Sharp and Dohme and TME Pharma.

Raoul Sutter received research grants from the Swiss National Foundation (No 320030_169379), the Research Fund of the University Basel, the Scientific Society Basel, and the Gottfried Julia Bangerter-Rhyner Foundation. He received personal grants from UCB-pharma and holds stocks from Alcon, Novartis, Roche, and Johnson & Johnson. Stefan Rüegg has received funding from the Swiss National Science Foundation (grant number 320030_169379/1). He has received honoraria from serving on the scientific advisory boards of Arvelle/Angelini, Bial, Eisai, GW, and UCB-pharma, and from serving as a consultant for Arvelle/Angelini, Eisai, Pfizer, Novartis, Sandoz, and UCB-pharma. He has received speaker’s honoraria from Bial, Eisai and Novartis. He does not hold any stocks of any pharmaceutical industries or manufacturers of medical devices. He disclosed that he is Editor-in-chief of Zeitschrift für Epileptologie/ Clinical Epileptology (01/01/2022; no payments).

Margitta Seeck has shares in Clouds of Care and dEEGtal, and received speaker honoraria from Bial and EISAI. She was funded by the Swiss National Science Foundation (No 180365). Matthew Walker has consulted for Angelini, EpilepsyGtX and Seer and has received honoraria from Angelini, Bioquest, Eisai and UCB pharma. Matthias Koepp or his department has received grants from Angelini, Biohavn, UCB and Xenon, has received consulting fees from Angelini, Bial, Biocodex, Eisai, Jazz Pharma, LivaNova, Sanofi and UCB Pharma, and is co-founder of PrevEp, Inc., and has received research funding from the MRC, Wellcome Trust, Epilepsy Research UK, Epilepsy Society UK, UCLH Foundation Trust, and Henry Smith Foundation. Eugen Trinka reports personal fees from Marinus, Angelini, Argenx, Alexion, Medtronic, BIAL – Portela & Cª, S.A, NewBridge, LivaNova, Eisai, UCB, Biogen, Sanofi, Jazz Pharmaceuticals, STOKE Therapeutics, Rapport, and Actavis. He is co-director of the European Consortium on Epilepsy Trials (ECET).

His institution received grants from Biogen, UCB Pharma, Eisai, Red Bull, Merck, Bayer, the European Union, FWF Österreichischer Fond zur Wissenschaftsforderung, Bundesministerium für Wissenschaft und Forschung, and Jubiläumsfond der Österreichischen Nationalbank. None related to the presented work. Moritz L Schmidbauer received grants from the Deutsche Forschungsgemeinschaft (DFG, TRR 274). Urs Fisch received research grants from the Bangerter-Rhyner Foundation and the Bouriez Foundation, none of which are related to this work. John Duncan reports research grants from National Institute for Health Research and Epilepsy Research Institute. MH served on scientific advisory boards of Biogen, Merck Serono, Alexion, Roche and Horizon Therapeutics (Amgen), received speaker’s honoraria from Biogen and received travel funding from Roche. Her institution received an unrestricted research grant from Roche. Rebecca Liu received personal fees from Angelini, Eisai and UCB Pharma and has received research funding from the Wellcome Trust. FLWVJS was supported by grants from the American Epilepsy Society (846534) and National Institutes of Health (R01NS127892).

All other authors do not report conflicts of interest.

## Data Availability

Data are available upon reasonable request with formal applications submitted to the respective cohort to protect patient sensitive data. Data from specific cohorts can be requested from the following cohort leaders: University Hospital Zurich, MG; University Hospital of Basel, UF; Hopitaux Universitaires de Geneve, PdS; University College London, SR; LMU University Hospital, MLS; Christian Doppler University Hospital Salzburg, GK; Kantonsspital St. Gallen, DZ; Prospective Regional Epilepsy Database and Biobank for Individualized Clinical Treatment, JZ. The ADNI data can be requested at the ADNI database (https://adni.loni.usc.edu/). Healthy control data from the Neuromorphometry by Computer Algorithm Chicago and Parkinson Progression Marker Initiative can be requested at https://schizconnect.org and https://www.ppmi-info.org respectively.

https://schizconnect.org

https://www.ppmi-info.org

## Acknowledgments

This study used data of people with Alzheimer’s disease and mild cognitive impairment from Alzheimer’s Disease Neuroimaging Initiative (ADNI) that was funded by National Institutes of Health Grant U01 AG024904 and DOD ADNI (Department of Defense award number W81XWH-12-2-0012). ADNI is funded by the National Institute on Aging, the National Institute of Biomedical Imaging and Bioengineering, and through generous contributions from the following: AbbVie; Alzheimer’s Association; Alzheimer’s Drug Discovery Foundation; Araclon Biotech; BioClinica, Inc.; Biogen; Bristol-Myers Squibb Company; CereSpir, Inc.; Cogstate; Eisai Inc.; Elan Pharmaceuticals, Inc.; Eli Lilly and Company; EuroImmun; F. Hoffmann-La Roche Ltd and its affiliated company Genentech, Inc.; Fujirebio; GE Healthcare; IXICO Ltd.; Janssen Alzheimer Immunotherapy Research & Development, LLC.; Johnson & Johnson Pharmaceutical Research & Development LLC.; Lumosity; Lundbeck; Merck & Co., Inc.; Meso Scale Diagnostics, LLC.; NeuroRx Research; Neurotrack Technologies; Novartis Pharmaceuticals Corporation; Pfizer Inc.; Piramal Imaging; Servier; Takeda Pharmaceutical Company; and Transition Therapeutics. The Canadian Institutes of Health Research is providing funds to support ADNI clinical sites in Canada. Private sector contributions are facilitated by the Foundation for the National Institutes of Health (www.fnih.org). The grantee organization is the Northern California Institute for Research and Education, and the study is coordinated by the Alzheimer’s Therapeutic Research Institute at the University of Southern California. ADNI data are disseminated by the Laboratory for Neuro Imaging at the University of Southern California. The PREDICT study is funded by the Swedish state through the ALF-agreement, Ann-Louise and Sven-Erik Biegler foundation, Hjärnfonden, Jeansson foundation, the Swedish Society for Medical Research, the Edit Jacobsson foundation, Rune and Ulla Amlöv Foundation.

---

**Extended Data Figure 1:**
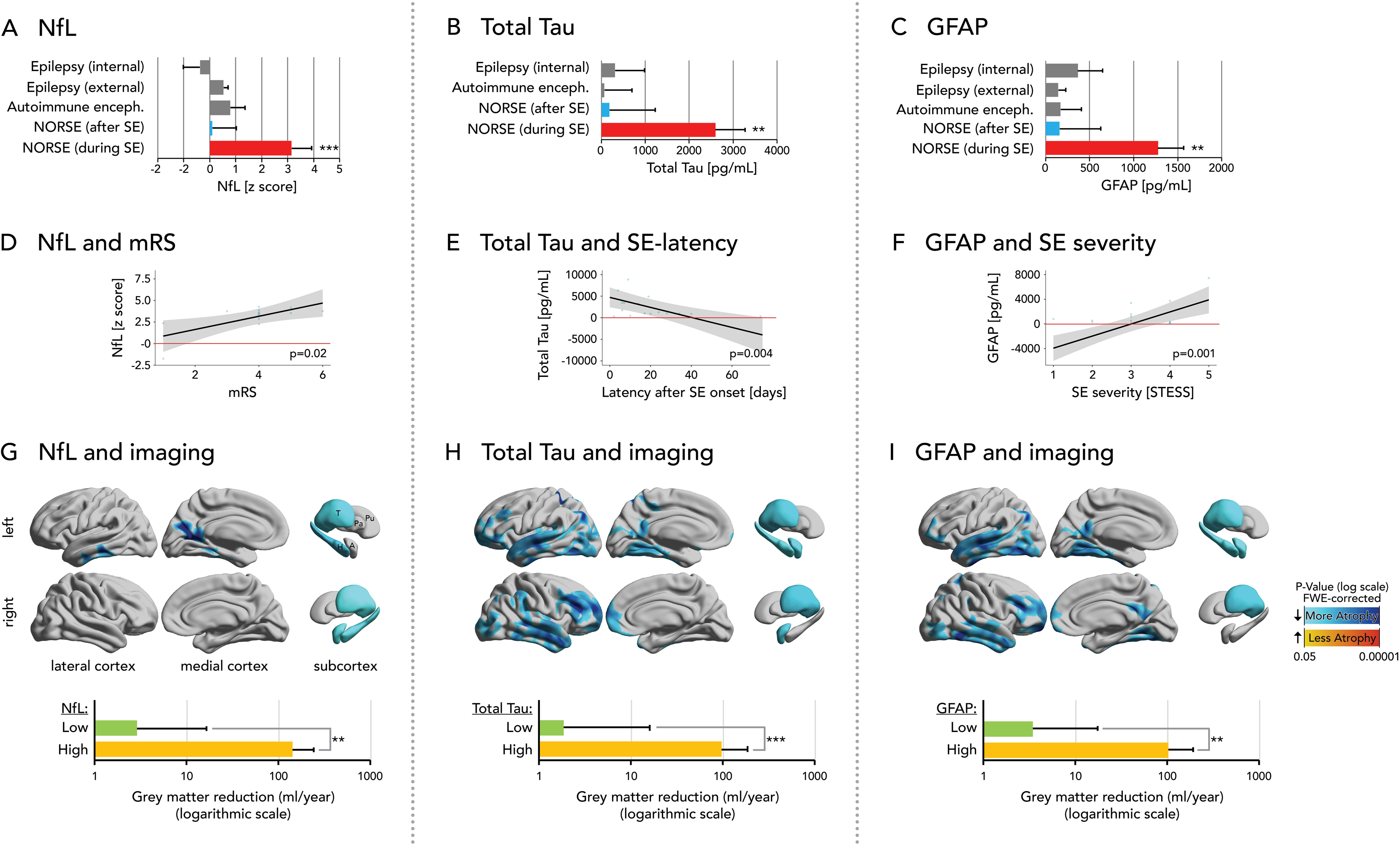
Grey matter volume loss in NORSE due to autoimmune encephalitis vs. autoimmune encephalitis without status epilepticus. Annualized grey matter volume (GMV) loss (ml/year, log scale) estimated using a linear mixed-effects model. GMV loss was approximately 18-fold higher in patients with NORSE due to autoimmune encephalitis (red) compared to those with autoimmune encephalitis without status epilepticus (grey). Horizontal lines indicate 95% confidence intervals (upper bounds shown).

**Extended Data Figure 2:**
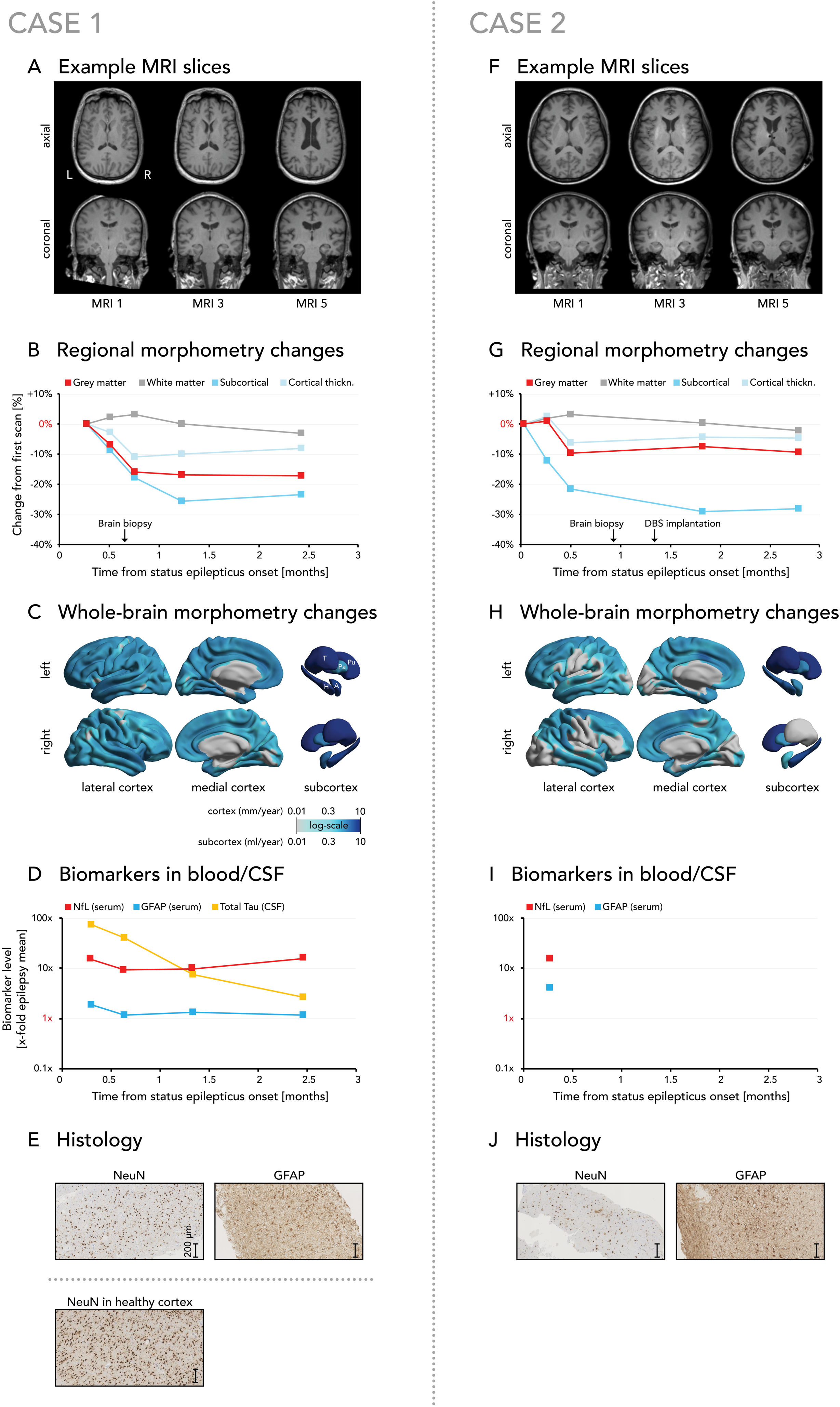
Distribution of bootstrapped t and p values. Density plots showing the distribution of p-values **(A)** and t-values **(B)** derived from 1,000 bootstrapped analysis samples for the analysis comparing the rate of grey matter volume reduction during NORSE with healthy controls using linear mixed effects models.

**Extended Data Table 1:** Baseline characteristics of individuals with NORSE.

|  | NORSE<br>Full Cohort<br>(n = 33) | NORSE<br>Discovery<br>Cohort<br>(n = 16) | NORSE<br>Replication Cohort<br>(n = 17) |
| --- | --- | --- | --- |
| Sex |  |  |  |
| Male | 17 (52%) | 7 (44%) | 10 (59%) |
| Female | 16 (48%) | 9 (56%) | 7 (41%) |
| Age |  |  |  |
| Age at baseline scan (*years*) | 50 ± 20 | 48 ± 18 | 52 ± 24 |
| Aetiology |  |  |  |
| Autoimmune encephalitis | 10 (30%) | 4 (25%) | 6 (35%) |
| Viral encephalitis | 7 (21%) | 3 (19%) | 4 (24%) |
| Other known aetiology | 3 (9%) | 1 (6%) | 2 (12%) |
| Cryptogenic | 13 (39%) | 8 (50%) | 5 (29%) |
| Clinical characteristics |  |  |  |
| Prodromal phase | 30 (91%) | 15 (94%) | 15 (88%) |
| Fever before onset | 14 (42%) | 7 (44%) | 7 (41%) |
| Status epilepticus severity (*STESS*) | 3.4 ± 1.5 | 3 ± 1.35 | 3.7 ± 1.5 |
| Duration of status epilepticus (*days*) | 30 ± 41 | 29 ± 43 | 31 ± 52 |
| Outcome |  |  |  |
| Duration ICU stay (*days*) | 44 ± 41 | 34 ± 40 | 54 ± 54 |
| Duration hospital stay (*days*) | 71 ± 77 | 52 ± 58 | 92 ± 104 |
| Disability at discharge (*mRS*) | 4.0 ± 1.4 | 4.3 ± 1.22 | 3.8 ± 1.6 |
| Death at last follow-up | 7 (21%) | 4 (25%) | 3 (18%) |
| Antiseizure medications |  |  |  |
| Number of antiseizure medications | 6 ± 2 | 6 ± 2 | 5 ± 2 |
| Levetiracetam | 31 (94%) | 16 (100%) | 15 (88%) |
| Phenytoin | 17 (52%) | 8 (50%) | 9 (53%) |
| Lacosamide | 27 (82%) | 14 (88%) | 13 (76%) |
| Phenobarbital | 21 (64%) | 13 (81%) | 8 (47%) |
| Sodium valproate | 21 (64%) | 11 (69%) | 10 (59%) |
| Lamotrigine | 3 (9%) | 0 (0%) | 3 (18%) |
| Perampanel | 11 (33%) | 7 (44%) | 4 (24%) |
| Brivaracetam | 6 (18%) | 4 (25%) | 2 (12%) |
| Gabapentin | 0 (0%) | 0 (0%) | 0 (0%) |
| Carbamazepine | 3 (9%) | 2 (13%) | 1 (6%) |
| Oxcarbazepine | 1 (3%) | 1 (6%) | 0 (0%) |
| Topiramate | 7 (21%) | 3 (19%) | 4 (24%) |
| Ketogenic diet | 5 (15%) | 1 (6%) | 4 (24%) |
| Anesthetic drugs |  |  |  |
| Number of anesthetic drugs | 2 ± 1 | 3 ± 1 | 2 ± 1 |
| Midazolam | 23 (70%) | 14 (88%) | 9 (53%) |
| Propofol | 20 (61%) | 8 (50%) | 12 (71%) |
| Ketamine | 14 (42%) | 11 (69%) | 3 (18%) |
| Thiopental | 13 (39%) | 9 (56%) | 4 (24%) |
| Inhalative anesthetics (iso/sevofluran) | 3 (9%) | 3 (19%) | 0 (0%) |
| Antiinfectious Therapy |  |  |  |
| Antibiotics | 25 (76%) | 11 (69%) | 14 (82%) |
| Antiviral Therapy | 23 (70%) | 10 (63%) | 13 (76%) |
| Immunotherapy |  |  |  |
| Steroids | 23 (70%) | 12 (75%) | 11 (65%) |
| IVIg | 9 (27%) | 2 (13%) | 7 (41%) |
| Plasma exchange | 10 (30%) | 6 (38%) | 4 (24%) |
| Rituximab | 8 (24%) | 2 (13%) | 6 (35%) |
| Anakinra | 3 (9%) | 0 | 2 (12%) |
| Tocilizumab | 1 (3%) | 0 | 1 (6%) |
| MRI scanning (*per scan*) |  |  |  |
| Number of scans | 90 | 48 | 42 |
| Field strength |  |  |  |
| 1.5 Tesla | 18 (20%) | 1 (2%) | 17 (40%) |
| 3 Tesla | 72 (80%) | 47 (98%) | 25 (60%) |
| Manufacturer |  |  |  |
| Siemens | 82 (91%) | 44 (92%) | 38 (90%) |
| General Electric | 5 (6%) | 3 (6%) | 2 (5%) |
| Philips | 3 (3%) | 1 (2%) | 2 (5%) |
| Interscan interval (*months*) | 3.5 ± 12.3 | 2.3 ± 12.7 | 5.6 ± 17.7 |
| Image quality rating | 1.9 ± 0.3 | 1.8 ± 0.3 | 2.0 ± 0.3 |
| Coefficient of variation |  |  |  |
| Cortical thickness | 0.06 | 0.06 | 0.07 |
| Grey matter volume | 0.1 | 0.1 | 0.11 |

Data presented as N (%) or as mean ± standard deviation. *NORSE*, new onset refractory status epilepticus; *ICU*, intensive care unit; *mRS*, modified Rankin scale; *IVIG*, intravenous immunoglobulins.

**Extended Data Table 2:** Baseline characteristics of control cohorts.

|  | Healthy controls<br>(n = 99) | Epilepsy with brief seizures<br>(n = 115) | Autoimmune Encephalitis<br>(n = 31) | Subarachnoid Hemorrhage<br>(n = 23) | Mild cognitive impairment<br>(n = 155) | Alzheimer's Disease<br>(n = 103) |
| --- | --- | --- | --- | --- | --- | --- |
| Sex |  |  |  |  |  |  |
| Male | 50 (51%) | 53 (46%) | 17 (55%) | 5 (22%) | 111 (72%) | 53 (51%) |
| Female | 49 (49%) | 62 (54%) | 14 (45%) | 18 (78%) | 44 (28%) | 50 (49%) |
| Age |  |  |  |  |  |  |
| Age at baseline scan | 47 ± 16 | 37 ± 15 | 49 ± 17 | 57 ± 16 | 75 ± 7 | 75 ± 8 |
| MRI scanning (*per scan*) |  |  |  |  |  |  |
| Number of scans | 198 | 230 | 75 | 58 | 958 | 446 |
| Field strength |  |  |  |  |  |  |
| 1.5 Tesla | 98 (49%) | 230 (100%) | 9 (12%) | 2 (3%) | 839 (88%) | 375 (84%) |
| 3 Tesla | 100 (51%) | 0 | 66 (88%) | 56 (97%) | 119 (12%) | 72 (16%) |
| Manufacturer |  |  |  |  |  |  |
| Siemens | 100 (51%) | 0 (0%) | 51 (68%) | 1 (2%) | 366 (38%) | 166 (37%) |
| General Electric | 98 (49%) | 230 (100%) | 10 (13%) | 7 (12%) | 532 (55%) | 209 (47%) |
| Philips | 0 (0%) | 0 (0%) | 14 (19%) | 50 (86%) | 60 (6%) | 71 (16%) |
| Interscan interval (*months*) | 29 ± 14 | 42 ± 3 | 9.6 ± 16.8 | 7.1 ± 23.1 | 7.2 ± 2.8 | 7.5 ± 3.7 |
| Image quality rating | 2.1 ± 0.2 | 2.2 ± 0.1 | 1.8 ± 0.1 | 1.8 ± 0.1 | 2.1 ± 0.2 | 2.1 ± 0.2 |
| Coefficient of variation |  |  |  |  |  |  |
| Cortical thickness | 0.06 | 0.05 | 0.07 | 0.05 | 0.06 | 0.07 |
| Grey matter volume | 0.13 | 0.14 | 0.12 | 0.11 | 0.1 | 0.11 |

Data presented as N (%) or as mean ± standard deviation.

**Extended Data Table 3:** Comparison of included NORSE cases with total NORSE cohort at the University Hospital Zurich.

|  | NORSE internal<br>included<br>(n = 16) | NORSE internal<br>total<br>(n = 28) | P value |
| --- | --- | --- | --- |
| Sex |  |  |  |
| Male | 7 (44%) | 11 (39%) | 1.00 |
| Female | 9 (56%) | 17 (61%) | 1.00 |
| Age |  |  |  |
| Age at baseline scan | 48 ± 18 | 54 ± 17 | 1.00 |
| Aetiology |  |  |  |
| Autoimmune encephalitis | 4 (25%) | 4 (14%) | 0.43 |
| Viral encephalitis | 3 (19%) | 5 (18%) | 1.00 |
| Other known aetiology | 1 (6%) | 2 (7%) | 1.00 |
| Cryptogenic | 8 (50%) | 19 (68%) | 0.34 |
| Clinical characteristics |  |  |  |
| Prodromal phase | 15 (94%) | 24 (86%) | 0.64 |
| Fever before onset | 7 (44%) | 15 (54%) | 0.75 |
| Status epilepticus severity (*STESS*) | 3 ± 1.35 | 3 ± 1.38 | 1.00 |
| Duration of status epilepticus (*days*) | 29 ± 43 | 25 ± 43 | 1.00 |
| Outcome |  |  |  |
| Duration ICU stay (*days*) | 34 ± 40 | 28 ± 39 | 1.00 |
| Duration hospital stay (*days*) | 52 ± 58 | 45 ± 57 | 1.00 |
| mRS at discharge | 4.3 ± 1.22 | 4.6 ± 1.26 | 1.00 |
| Death | 4 (25%) | 12 (43%) | 0.33 |
| Antiseizure medications |  |  |  |
| Number of antiseizure medications. | 6 ± 2 | 6 ± 3 | 1.00 |
| Levetiracetam | 16 (100%) | 27 (96%) | 1.00 |
| Phenytoin | 8 (50%) | 13 (46%) | 1.00 |
| Lacosamide | 14 (88%) | 18 (64%) | 0.16 |
| Phenobarbital | 13 (81%) | 21 (75%) | 0.72 |
| Sodium valproate | 11 (69%) | 20 (71%) | 1.00 |
| Lamotrigine | 0 (0%) | 1 (4%) | 1.00 |
| Perampanel | 7 (44%) | 10 (36%) | 0.75 |
| Brivaracetam | 4 (25%) | 5 (18%) | 0.7 |
| Gabapentin | 0 (0%) | 1 (4%) | 1.00 |
| Carbamazepine | 2 (13%) | 2 (7%) | 0.61 |
| Oxcarbazepine | 1 (6%) | 2 (7%) | 1.00 |
| Topiramate | 3 (19%) | 4 (14%) | 0.69 |
| Clonazepam/Lorazepam | 5 (31%) | 11 (39%) | 0.75 |
| Anesthetics |  |  |  |
| Number of anesthetics | 3 ± 1 | 2 ± 1 | 1.00 |
| Midazolam | 14 (88%) | 24 (86%) | 1.00 |
| Propofol | 8 (50%) | 12 (43%) | 0.76 |
| Ketamine | 11 (69%) | 14 (50%) | 0.34 |
| Thiopental | 9 (56%) | 15 (54%) | 1.00 |
| Inhalative anesthetics | 3 (19%) | 3 (11%) | 0.65 |
| Antiinfectious Therapy |  |  |  |
| Antibiotics | 11 (69%) | 17 (61%) | 0.75 |
| Antiviral Therapy | 10 (63%) | 17 (61%) | 1.00 |
| Immunotherapy |  |  |  |
| Steroids | 12 (75%) | 17 (61%) | 0.51 |
| IVIg | 2 (13%) | 2 (7%) | 0.61 |
| Plasma exchange | 6 (38%) | 8 (29%) | 0.74 |
| Rituximab | 2 (13%) | 3 (11%) | 1.00 |
| Anakinra | 0 | 0 | 1.00 |
| Tocilizumab | 0 | 0 | 1.00 |
| Ketogenic diet | 1 (6%) | 1 (4%) | 1.00 |

Data presented as N (%) and analysed with Fisher's exact test or as mean ± standard deviation and analysed with the Mann-Whitney U test.
NORSE, new onset refractory status epilepticus; ICU, intensive care unit; mRS, modified Rankin scale; IVIG, intravenous immunoglobulins.

**Extended Data Table 4:** Secondary analyses for longitudinal brain atrophy in NORSE vs. healthy aging.

| Secondary analysis | GMV reduction (mL/year) |  | N |  | T-value | P-value |
| --- | --- | --- | --- | --- | --- | --- |
|  | NORSE | Healthy controls | NORSE | Healthy controls |  |  |
| FULL ANALYSIS | 234.3 ± 66.2 | 3.0 ± 1.8 | 33 | 99 | 6.85 | <0.001 |
| Leave-one-cohort-out |  |  |  |  |  |  |
| Basel left out | 249.8 ± 65.6 | 3.0 ± 1.7 | 28 | 99 | 7.38 | <0.001 |
| Geneva left out | 236.1 ± 66.3 | 3.0 ± 1.8 | 32 | 99 | 6.90 | <0.001 |
| London left out | 235.5 ± 66.9 | 3.0 ± 1.8 | 31 | 99 | 6.82 | <0.001 |
| Munich left out | 294.7 ± 73.8 | 3.0 ± 1.7 | 28 | 99 | 7.75 | <0.001 |
| Salzburg left out | 217.8 ± 63.8 | 3.0 ± 1.7 | 30 | 99 | 6.60 | <0.001 |
| St. Gallen left out | 220.8 ± 67.9 | 3.0 ± 1.7 | 32 | 99 | 6.29 | <0.001 |
| Zurich left out | 147.2 ± 86.1 | 3.0 ± 1.3 | 17 | 99 | 3.28 | 0.001 |
| Healthy: London left out | 223.8 ± 73.2 | 3.2 ± 5.5 | 33 | 50 | 5.93 | <0.001 |
| Healthy: NMorph left out | 233.5 ± 66.8 | 2.8 ± 1.8 | 33 | 87 | 6.78 | <0.001 |
| Healthy: PPMI left out | 233.9 ± 75.3 | 3.1 ± 2.1 | 33 | 61 | 6.01 | <0.001 |
| Scanner aspects |  |  |  |  |  |  |
| Only re-scans on same scanner | 239.1 ± 61.8 | 3.1 ± 1.5 | 26 | 99 | 7.49 | <0.001 |
| Only 3T MRI scans | 296.1 ± 83.1 | 3.2 ± 5.4 | 28 | 50 | 6.92 | <0.001 |
| Only Siemens MRI scans | 235.2 ± 82.1 | 3.2 ± 5.5 | 32 | 50 | 5.55 | <0.001 |

A set of secondary analyses was conducted to assess the robustness of the main findings, shown on top. First, we performed leave-one-cohort-out cross-validation by repeating the analysis in subcohorts with each reported cohort excluded in turn. Second, we evaluated the impact of scanner-related factors by (i) restricting analyses to scans and rescans performed on the same scanner, (ii) limiting the dataset to 3T MRI scans, and (iii) including only scans acquired on Siemens MRI systems. All statistical analyses mirrored the primary analysis, comparing GMV reduction rates in NORSE vs. healthy controls using linear mixed-effects models adjusted for age at first scan, interscan interval, sex, baseline group differences, and total intracranial volume, with subject as a random effect to account for repeated measures.
GMV, grey matter volume; N, number; NMorph, Neuromorphometry by Computer Algorithm Chicago; PPMI, Parkinson's Progression Markers Initiative, 3T, 3 Tesla.

**Extended Data Table 5:**
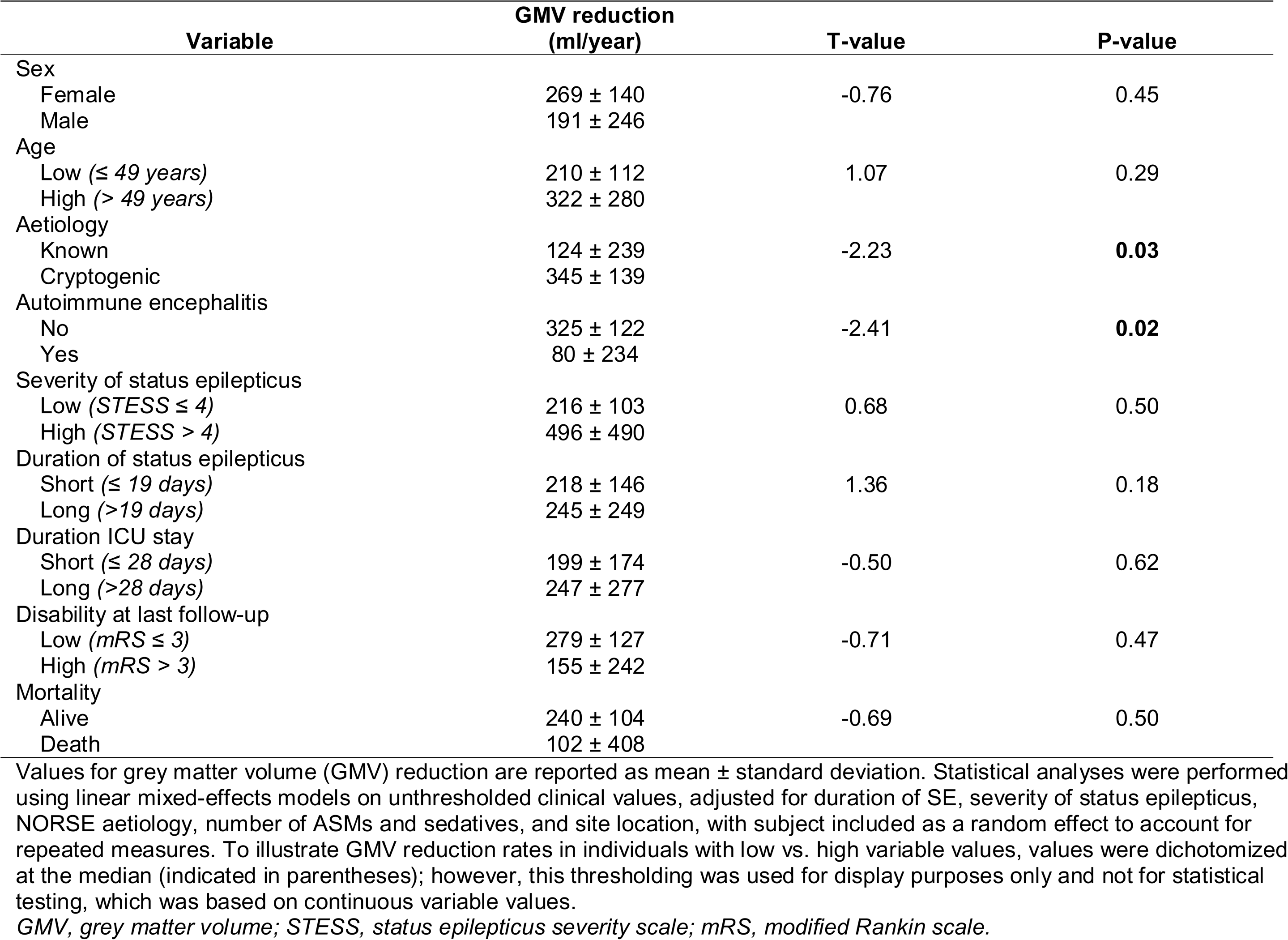
Clinical variables and their association with the rate of grey matter volume reduction in NORSE.

| Variable | GMV reduction (ml/year) | T-value | P-value |
| --- | --- | --- | --- |
| Sex |  |  |  |
| Female | 269 ± 140 | -0.76 | 0.45 |
| Male | 191 ± 246 |  |  |
| Age |  |  |  |
| Low ($\leq 49$ years) | 210 ± 112 | 1.07 | 0.29 |
| High ($> 49$ years) | 322 ± 280 | | |
| Aetiology |  |  |  |
| Known | 124 ± 239 | -2.23 | **0.03** |
| Cryptogenic | 345 ± 139 |  |  |
| Autoimmune encephalitis |  |  |  |
| No | 325 ± 122 | -2.41 | **0.02** |
| Yes | 80 ± 234 |  |  |
| Severity of status epilepticus |  |  |  |
| Low ($STE\!SS \leq 4$) | 216 ± 103 | 0.68 | 0.50 |
| High ($STE\!SS > 4$) | 496 ± 490 | | |
| Duration of status epilepticus |  |  |  |
| Short ($\leq 19$ days) | 218 ± 146 | 1.36 | 0.18 |
| Long ($> 19$ days) | 245 ± 249 | | |
| Duration ICU stay |  |  |  |
| Short ($\leq 28$ days) | 199 ± 174 | -0.50 | 0.62 |
| Long ($> 28$ days) | 247 ± 277 | | |
| Disability at last follow-up |  |  |  |
| Low ($mRS \leq 3$) | 279 ± 127 | -0.71 | 0.47 |
| High ($mRS > 3$) | 155 ± 242 | | |
| Mortality |  |  |  |
| Alive | 240 ± 104 | -0.69 | 0.50 |
| Death | 102 ± 408 |  |  |

Values for grey matter volume (GMV) reduction are reported as mean ± standard deviation. Statistical analyses were performed using linear mixed-effects models on unthresholded clinical values, adjusted for duration of SE, severity of status epilepticus, NORSE aetiology, number of ASMs and sedatives, and site location, with subject included as a random effect to account for repeated measures. To illustrate GMV reduction rates in individuals with low vs. high variable values, values were dichotomized at the median (indicated in parentheses); however, this thresholding was used for display purposes only and not for statistical testing, which was based on continuous variable values.
GMV, grey matter volume; STESS, status epilepticus severity scale; mRS, modified Rankin scale.

**Extended Data Table 6:** Fluid biomarkers.

|  | NORSE<br>(during SE) | NORSE<br>(after SE) | Epilepsy<br>(internal) | Epilepsy<br>(external) | Autoimmune<br>encephalitis |
| --- | --- | --- | --- | --- | --- |
| **NfL** |  |  |  |  |  |
| Z score | 3.2 | 0.0 | -0.4 | 0.5 | 0.8 |
| 95% CI | (2.4–3.9) | (-1.0–1.0) | (-1.0–0.3) | (0.4–0.7) | (0.2–1.4) |
| Value (pg/mL) | 531 | 369 | 22 | 14 | 25 |
| 95% CI (pg/mL) | (423–638) | (254–483) | (-68–113) | (-11–39) | (-66–98) |
| N | 12 | 6 | 17 | 214 | 25 |
| T value | Ref. | 5.7 | 7.0 | 6.5 | 4.9 |
| P value | Ref. | <0.001 | <0.001 | <0.001 | <0.001 |
| **Total Tau** |  |  |  |  |  |
| Value (pg/mL) | 2599 | 182 | 313 | NA | 66 |
| 95% CI (pg/mL) | (1921–3277) | (-858–1221) | (-357–983) | NA | (-560–690) |
| N | 9 | 6 | 17 | 0 | 24 |
| T value | Ref. | 3.4 | 4.5 | NA | 4.8 |
| P value | Ref. | 0.007 | 0.004 | NA | <0.001 |
| **GFAP** |  |  |  |  |  |
| Value (pg/mL) | 1275 | 160 | 367 | 145 | 171 |
| 95% CI (pg/mL) | (986–1565) | (-306–626) | (85–648) | (61–230) | (-62–405) |
| N | 12 | 6 | 17 | 183 | 25 |
| T value | Ref. | 3.8 | 4.3 | 6.8 | 5.5 |
| P value | Ref. | 0.002 | <0.001 | <0.001 | <0.001 |

Values and z-scores are presented as mean with 95% confidence intervals. The smaller, internal epilepsy cohort was enrolled at University Hospital Zurich using the same sampling and processing protocols as the other study groups, whereas the larger, external cohort was drawn from an earlier regional biobank study in Västra Götaland, Sweden. Total Tau in cerebrospinal fluid was not available in the external epilepsy cohort. Statistical analyses were conducted using linear mixed-effects models, adjusted for age, sex, and body mass index, with subject as a random effect to account for repeated measures. T and P values reflect comparisons between each cohort and the NORSE group during status epilepticus, with Bonferroni correction applied for multiple testing.
NfL, neurofilament light chain; GFAP, glial fibrillary acidic protein; NORSE, new-onset refractory status epilepticus; SE, status epilepticus; CI, confidence interval.

**Extended Data Table 7:** Association of fluid biomarkers with the rate of GMV reduction.

| Variable | GMV reduction<br>(ml/year) | T-value | P-value |
| --- | --- | --- | --- |
| NfL |  |  |  |
| Low ($\leq 21$ /mL) | 3 ± 13 | 1.4 | **0.009** |
| High ($> 21$ pg/mL) | 141 ± 99 | | |
| Total-Tau |  |  |  |
| Low ($\leq 268$ pg/mL) | 2 ± 14 | 4.0 | **<0.001** |
| High ($> 268$ pg/mL) | 96 ± 88 | | |
| GFAP |  |  |  |
| Low ($\leq 196$ pg/mL) | 3 ± 14 | 3.0 | **0.004** |
| High ($> 196$ pg/mL) | 101 ± 90 | | |

Values for GMV reduction are reported as mean ± standard deviation. Statistical analyses were performed using linear mixed-effects models on unthresholded biomarker values, adjusted for age, sex, and body mass index, with subject included as a random effect to account for repeated measures. To illustrate GMV reduction rates in individuals with low vs. high biomarker levels, values were dichotomized at the median (indicated in parentheses); however, this thresholding was used for display purposes only and not for statistical testing, which was based on continuous biomarker values.
*NfL*, neurofilament light chain; *GFAP*, glial fibrillary acidic protein; *GMV*, grey matter volume.

## Notes

### Author Declarations

Ethics committees of the Canton of Zurich, Canton of Geneva and Canton of Basel gave ethical approval for the Swiss cohort for this work. Ethics committee for the Region of Salzburg gave ethical approval for the Austrian cohort for this work. Ethics committee for LMU Munich gave ethical approval for the German cohort for this work. For the UK cohort, the work was performed as part of a service evaluation, registered and independently approved by the Clinical Audit and Quality Improvement Subcommittee at UCLH University College London Hospitals Trust. This waives the need for approval by an ethics committee,

