## Supplementary information for "Brain Damage During New-Onset Refractory Status Epilepticus"

**Content:**

**1. Description of NORSE replication cohorts**

**1.1 Switzerland**

The Basel (Switzerland) cohort included consecutive adult patients (i.e., patients ≥18 years of age) diagnosed with new onset refractory status epilepticus (NORSE) between 2005 and 2023 who had been treated at the University Hospital of Basel, based on a local prospective status epilepticus (SE) registry. For this study, patients with NORSE having at least two longitudinal MRI scans at least 1 week apart, at least one of which was done during NORSE, were included. Patients younger than 18 years and those with a known neurodegenerative condition or mass lesion were excluded. Patients with repetitive seizures not qualifying as SE were excluded.

The Geneva (Switzerland) cohort included consecutive patients ≥16 years of age with diagnosed new onset status epilepticus at the University Hospital of Geneva based on a local prospective SE registry. For this study, those who had had at least two longitudinal MRI scans at least 1 week apart, at least one of which was done during NORSE, were included. Patients with a known neurodegenerative condition or a mass lesion that could have disrupted MR preprocessing were excluded.

The St. Gallen (Switzerland) cohort included patients ≥18 years of age with diagnosed new onset status epilepticus at the Cantonal Hospital of St. Gallen who had had at least two longitudinal MRI scans at least 1 week apart, at least one of which was done during NORSE. Patients with a known neurodegenerative condition or a mass lesion that could have disrupted MR preprocessing were excluded.

**1.2 Austria**

The Salzburg (Austria) cohort included consecutive adult patients (i.e., patients ≥18 years of age) with suspected or definitive SE between February 2019 and October 2023 in a prospective single-centre longitudinal MRI study to assess the contribution of peri-ictal MRI abnormalities to the outcome of SE (Bosque Varela et al., Epilepsy Behav 2024). For the current analysis, cases with confirmed new onset refractory status epilepticus without a history of active epilepsy or pre-existing neurological disorders were included if they received more than 2 longitudinal MRI scans during SE at least 1 week apart. Patients with clear acute or subacute structural, toxic or metabolic causes were excluded.

**1.3 Germany**

The Munich (Germany) cohort included consecutive adult patients (i.e., patients ≥18 years of age) recruited as a subset of a study concerned with new onset refractory status epilepticus which evolved into super refractory status epilepticus and enteral or parenteral phenobarbital treatment between September 30^th^ 2015 and September 30^th^ 2020 who had been treated at the hospitals associated with the Ludwig Maximilian University (LMU) in Munich. Patients with anoxic brain injury and patients in whom further antiseizure medications were added after the first administration of phenobarbital were excluded from the Munich cohort. NORSE cases having at least two longitudinal MRI scans at least 1 week apart, at least one of which was done during NORSE, were included.

**1.4 United Kingdom**

The UK cohort included patients recruited through a retrospective review of all patients admitted to the neurological intensive care unit (ICU) at the National Hospital for Neurology and Neurosurgery (NHNN) Queen Square, between January 2004 and September 2018 for SE with supra-refractory status epilepticus (SRSE). Of these the patients with NORSE were selected, as described previously (Neligan et al., Epilepsy Behav 2021). Inclusion criteria were defined as patients with new-onset refractory SE without a prior history of epilepsy, and in whom no clear aetiology was identified within 48 hours of admission. For this study, patients with NORSE having at least two longitudinal MRI scans at least 1 week apart, at least one of which was done during NORSE, were included. Patients younger than 18 years and those with a known neurodegenerative condition or mass lesion were excluded.

**1.5 Informed consent procedures**

Regulatory approval was granted by the local or regional ethical commissions. All subjects in the Swiss, Austrian, and German replication cohorts gave informed consent or consent was provided by a patient’s legal representative. For the UK cohort, the work was performed as part of a service evaluation, registered and independently approved by the Clinical Audit and Quality Improvement Subcommittee at UCLH University College London Hospitals Trust. This waives the need for approval by an ethics committee, and individual informed consent in accordance with UK legislation and NHS operating procedures.

**2. Description of comparison cohorts**

**2.1 Alzheimer’s Disease Neuroimaging Initiative (ADNI)**

To compare the rate of brain atrophy and neurodegeneration during NORSE to common neurodegenerative conditions, we included data of people with Alzheimer’s disease and mild cognitive impairment from ADNI.

**Table 1:** *Neurocognitive comparison cohort demographics and clinical characteristics*

|  | Mild cognitive impairment  (n = 155) | Dementia/Alzheimer’s Disease  (n = 103) |
| --- | --- | --- |
| **Sex** |  |  |
| **Female** | 44 (28%) | 50 (50%) |
| **Male** | 111 (72%) | 53 (50%) |
| **Age at baseline scan *(years)*** | 75 ± 7 | 75 ± 8 |
| **Interval between scans *(months)*** | 7.5 ± 3.7 | 7.2 ± 2.8 |
| **MMS** | 27 ± 2 | 23 ± 2 |
| **CDR** | 0.48 ± 0.1 | 0.74 ± 0.29 |
| **APO-E genotype** |  |  |
| **2/3** | 8 (5%) | 3 (3%) |
| **2/4** | 7 (5%) | 3 (3%) |
| **3/3** | 62 (40%) | 30 (29%) |
| **3/4** | 60 (39%) | 44 (43%) |
| **4/4** | 18 (12%) | 23 (22%) |

Obtained through: The data was obtained from the Alzheimer’s Disease Neuroimaging Initiative (ADNI), a longitudinal, non-interventional study designed to develop imaging, fluid and cognitive biomarkers of Alzheimer’s disease progression. Between 2003 and 2008 the original ADNI-1 phase recruited 800 participants (200 cognitively normal controls, 400 individuals with amnestic mild cognitive impairment [MCI] and 200 with mild Alzheimer’s disease [AD]) across ~50 academic sites in the United States and Canada. Key eligibility criteria were age 55–90 years, Hachinski Ischaemic Score ≤ 4, Geriatric Depression Scale < 6, availability of a study partner, and the absence of significant medical, psychiatric or neurological comorbidity. Diagnostic-group specific criteria required an MMSE of 24–30 for controls and MCI, 20–26 for AD, and a Clinical Dementia Rating (CDR) of 0 for controls, 0.5 for MCI and 0.5–1.0 for AD. All participants agreed to longitudinal follow-up and neuroimaging; 20–50 % also consented to serial lumbar punctures.

The ADNI was launched in 2003 as a public-private partnership, led by Principal Investigator Michael W. Weiner, MD. The primary goal of ADNI has been to test whether serial magnetic resonance imaging (MRI), positron emission tomography (PET), other biological markers, and clinical and neuropsychological assessment can be combined to measure the progression of mild cognitive impairment (MCI) and early Alzheimer’s disease (AD).

For the current analyses we downloaded all available T1-weighted scans for AD and MCI participants in ADNI-1, downloaded on 9^th^ August 2024.

Reference for dataset: adni.loni.usc.edu.

Imaging schedule and quality control: AD subjects were scanned at baseline, 6, 12 and 24 months; MCI and control participants at baseline, 6, 12, 24 and 36 months. Every site underwent rigorous MRI certification that required phantom and volunteer scans to demonstrate protocol fidelity before enrolment could begin; phantom imaging is repeated with each participant scan to monitor scanner stability. Raw DICOM data are uploaded to the Laboratory of Neuroimaging, where the Mayo Clinic MRI core performs central quality control for protocol compliance, artefacts and clinically relevant findings. Scans failing quality control or showing major incidental pathology are repeated or excluded.

MRI acquisition protocol: Structural images are acquired with a sagittal 3-dimensional magnetisation-prepared rapid gradient-echo (MPRAGE) sequence on 1.5 T scanners using the following target parameters: repetition time (TR) ≈ 2,400 ms, echo time (TE) ≈ 3–4 ms, inversion time (TI) ≈ 1,000 ms, flip angle ≈ 8°, field-of-view 24 cm, 170 contiguous slices, voxel size ~1 × 1 × 1.2 mm. Sites with 3 T capability repeat the same geometry at higher field strength to enable harmonised cross-field analyses. Full parameter tables are available in the publicly accessible ADNI MRI technical manual: https://adni.loni.usc.edu/wp-content/themes/freshnews-dev-v2/documents/mri/ADNI_MRI_overview_2.6.18.pdf

**2.3 Epilepsy with brief seizures**

To compare the rates of atrophy during NORSE with a cohort of people with epilepsy, we included a well-characterised, longitudinal population-based cohort. This cohort provides a benchmark for brain-volume trajectories in people with brief, self-limited seizures that do not have SE.

**Table 2:** *Epilepsy with brief seizures cohort demographics and clinical characteristics*

|  | **Focal epilepsy**  **(n = 85)** | **Generalised epilepsy (n = 31)** |
| --- | --- | --- |
| **Sex** |  |  |
| **Male** | 42 (49%) | 11 (36%) |
| **Female** | 43 (51%) | 20 (65%) |
| **Age & duration *(years)*** |  |  |
| **Age at baseline scan** | 39 ± 15 | 32 ± 14 |
| **Age at first afebrile seizure** | 21 ± 14 | 11 ± 6 |
| **Duration of epilepsy** | 17 ± 14 | 20 ± 15 |
| **Imaging** |  |  |
| **Lesional (incl. HS)** | 36 (42%) | 4 (13%) |
| **HS** | 17 (20%) | 0 (0%) |
| **Epilepsy localisation** |  |  |
| **Temporal lobe** | 51 (60%) | 0 (0%) |
| **Frontal lobe** | 8 (9%) | 0 (0%) |
| **Generalised** | 0 (0%) | 31 (100%) |
| **Unknown / Undetermined** | 26 (31%) | 0 (0%) |
| **Seizures** |  |  |
| **Seizure frequency (any type, *per year*)** | 211 ± 567 | 604 ± 1319 |
| **Seizure frequency (any seizures, *categoric.*)** |  |  |
| **No seizures** | 22 (27%) | 4 (13%) |
| **Less than once a month** | 17 (21%) | 5 (17%) |
| **Once a month to once a week** | 16 (20%) | 6 (20%) |
| **Once a week to daily** | 26 (32%) | 15 (50%) |
| **Daily seizures** | 0 (0%) | 0 (0%) |
| **GTCS** |  |  |
| **History of GTCS** | 52/81 (64%) | 25/30 (83%) |
| **GTCS in last 6 months** | 34/81 (42%) | 21/30 (70%) |
| **GTCS frequency *(per year)*** | 18 ± 59 | 24 ± 49 |
| **No. of antiseizure medications at baseline** | 1.5 ± 1.0 | 1.5 ± 0.9 |

Obtained through: The control epilepsy group was drawn from a prospective, population-based MRI study of adults with chronic active epilepsy or newly diagnosed epilepsy living within a 15-mile radius of the Chalfont Centre for Epilepsy, Buckinghamshire, UK. Case ascertainment between June 1995 and May 1997 relied on systematic searches of 21 general-practice registers covering 207,553 residents, followed by expert electro-clinical adjudication in accordance with International League Against Epilepsy (ILAE) criteria.

For the present analysis we retained only participants with definitive epilepsy meeting the current practical ILAE definition of epilepsy, either newly-diagnosed or chronic, and who contributed ≥ 2 usable MRI examinations. We excluded participants with single seizures not meeting the criteria for epilepsy, acute-symptomatic seizures, or status epilepticus.

Recruitment and follow-up procedures were community-based: patients were contacted via their general practitioners, issued seizure diaries, and revisited 3.5 years later; attrition was minimised through national health-register tracing and accounted for refusals, death, emigration, or loss to follow-up (overall attrition 11 %). Participants with imaging evidence of cerebrovascular disease or other progressive structural lesions were excluded at baseline. The study received approval from the Joint Research Ethics Committee of the National Hospital for Neurology & Neurosurgery and University College London, and all subjects gave written informed consent.

Reference manuscript: *Liu, R. S. N. et al. A Longitudinal Quantitative MRI Study of Community-Based Patients with Chronic Epilepsy and Newly Diagnosed Seizures: Methodology and Preliminary Findings. NeuroImage 14, 231–243 (2001).*

MRI acquisition and quality control: Baseline and follow-up scans were obtained on the same 1.5 T GE Signa Horizon Echospeed scanner using an harmonised protocol:

- Sagittal 3-D inversion-recovery–prepared SPGR (IR-SPGR) T1-weighted volume: TR/TE/TI 17.4/4.2/450 ms, flip angle 20°, matrix 256 × 192, field-of-view 24 × 18 cm, 124 contiguous coronal slices, 1.5 mm thickness.
- Oblique-coronal proton-density and T2 spin-echo: TR 2000 ms / TE 30 ms (PD) and 120 ms (T2).
- Oblique-coronal FLAIR: TR/TE/TI 11 000/2600/144 ms, echo-train length 8.

Scans exhibiting severe motion were discarded. Longitudinal pairs were visually reviewed by two blinded consultant neuroradiologists for new lesions or surgery.

**2.4 Subarachnoid haemorrhage**

Prolonged ICU stays and exposure to potentially neurotoxic treatments may influence the rate of brain atrophy and neurodegeneration. To assess these effects independently of NORSE, we included a comparison cohort of individuals with subarachnoid haemorrhage (SAH), who had a median ICU stay of 24 days, as detailed below.

**Table 3:** *Subarachnoid haemorrhage* *cohort demographics and clinical characteristics*

| **Variable** | N (%) or  Median ± IQR |
| --- | --- |
| **Sex** |  |
| **Female** | 18 (78%) |
| **Male** | 5 (22%) |
| **Age at baseline scan *(years)*** | 57 ± 16 |
| **Interval between scans *(years)***  **Days on ICU**  **Days in hospital**  **Death during hospitalisation**  **Grading/classification**  **Hunt & Hess classification**  **WFNS scale**  **Fisher scale** | 7.1 ± 23.1  24 ± 15  31 ± 14  3 (23%)  2 ± 1  2 ± 1  4 ± 1 |

Obtained through: The SAH cohort included adult patients with a diagnosed aneurysmatic SAH who were admitted at the neurocritical care unit of the University Hospital Zurich between January 01^st^ 2017 and August 31^st^ 2024. We included them in the study if they had at least 2 high-resolution T1-weighted anatomical MRI images acquired with at least a 1 week interval between the images. We excluded participants with SE, neurodegenerative conditions or large mass lesions that may disrupt MRI processing.

Informed consent procedures: All subjects in the SAH cohort or their legal representatives gave written informed consent for the retrospective analysis of routinely collected data. The Zurich regional ethical committee approved the inclusion of the patients.

**2.5 Autoimmune encephalitis**

Neuroinflammation may impact the rate of brain atrophy and neurodegeneration. To assess these effects independently of NORSE, we included a comparison cohort of individuals with active definite autoimmune encephalitis, as detailed below.

Obtained through: The active autoimmune encephalitis cohort included adult patients with a definite autoimmune encephalitis according to established criteria (Graus et al., Lancet Neurol 2017) who were treated at the University Hospital Zurich between January 2013 and December 2024. Disease activity was categorized as “active” in patients suffering from an acute disease, a relapse or a chronic progressive disease course. Participants were included in the study if they had at least 2 high-resolution T1-weighted anatomical MRI images acquired with at least a 1 week interval between the images. We excluded participants with current or previous SE, neurodegenerative conditions or large mass lesions that may disrupt MRI processing.

**Table 4:** *Autoimmune encephalitis cohort demographics and clinical characteristics*

|  | N (%) or  Median ± IQR |
| --- | --- |
| **Sex** |  |
| **Female** | 18 (55%) |
| **Male** | 15 (45%) |
| **Age at baseline scan *(years)*** | 49 ± 17 |
| **Interval between scans *(years)***  **Subtype Encephalitis**  **Anti-NMDA-R**  **Anti-LG1**  **Anti-GAD**  **Anti-CASPR2**  **Anti-Gaba-AR**  **Anti-Gly-R**  **Anti-GABA-AR/Anti-GAD-AR**  **Anti-Hu**  **Anti-Ma**  **Anti-IgLON5**  **Anti-DPPX**  **Seronegative**  **Diagnosed tumor**  **Acute seizures** | 9.6 ± 16.8  3 (10%)  8 (26%)  3 (10%)  2 (6%)  3 (10%)  1 (3%)  1 (3%)  1 (3%)  1 (3%)  1 (3%)  1 (3%)  6 (19%)  5 (16%)  25 (80%) |

Informed consent procedures: All subjects in the autoimmune encephalitis cohort or their legal representatives gave written informed consent for the retrospective analysis of routinely collected data. The Zurich regional ethical committee approved the inclusion of the patients.

**3. Description of healthy control cohorts**

We included data of healthy volunteers from three cohorts: The NMorphCH study, the PPMI group and a prospective longitudinal MRI study in epilepsy and healthy volunteers (PNDE). By including these three cohorts we ensured that the data is representative and provides a diverse set of scanning protocols. We used propensity score matching (see below) to match the healthy volunteer data with the NORSE data for age and sex, as described below.

**Table 5:** *Healthy control demographics by cohort*

|  | NMorphCH  (n = 12) | PPMI  (n = 38) | PNDE  (n = 49) |
| --- | --- | --- | --- |
| **Sex** |  |  |  |
| **Female** | 5 (42%) | 19 (50%) | 25 (51%) |
| **Male** | 7 (58%) | 19 (50%) | 24 (49%) |
| **Age at baseline scan *(years)*** | 34 ± 9 | 58 ± 9 | 41 ± 16 |
| **Interval between scans *(years)*** | 1.65 ± 0.24 | 1.13 ± 0.31 | 3.53 ± 0.09 |

**3.1 Neuromorphometry by Computer Algorithm Chicago (NMorphCH)**

Number of volunteers: 12

Reference for dataset: <http://nunda.northwestern.edu/nunda/data/projects/NMorphCH>

Obtained through: ShizConnect

Reference for ShizConnect: *Kogan A, Alpert K, Ambite JL, Marcus DS, Wang L. Northwestern University schizophrenia data sharing for SchizConnect: A longitudinal dataset for large-scale integration. Neuroimage 2016; 124: 1196–201.*

MR-acquisition: 3T Siemens MRI scanner (Siemens Medical, Erlangen, Germany). A magnetization-prepared rapid gradient echo (MPRAGE) sequence was used to acquire high-resolution T1-weighted anatomical images (repetition time=2400 ms, echo time=3.16 ms, flip=8°, 256 x 256 matrix, 176 slices, slice thickness=1.0 mm, voxel size=1x1x1mm^3^)

**3.2 Parkinson Progression Marker Initiative (PPMI)**

Number of volunteers: 38

Reference for dataset: *Parkinson Progression Marker Initiative. The Parkinson Progression Marker Initiative (PPMI). Progress in Neurobiology 2011; 95: 629-35.*

MR-acquisition: Scans used for this longitudinal cohort were acquired on 3T Siemens MRI scanners (Siemens Medical, Erlangen, Germany). A magnetization-prepared rapid gradient echo (MPRAGE) sequence was used to acquire high-resolution T1-weighted anatomical images (repetition time=2300 ms, echo time=2.98 ms, flip angle=9°, 240 x 256 matrix, 160-192 slices, slice thickness=1.0 mm, voxel size=1x1x1mm^3^). The T1 acquisition protocol followed ADNI-3 sequence parameter recommendations: <http://adni.loni.usc.edu/wp-content/uploads/2017/07/ADNI3-MRI-protocols.pdf>

Detailed description of MR-acquisition protocol can be found in the PPMI MRI Technical Operations Manual: <http://www.ppmi-info.org/wp-content/uploads/2017/06/PPMI-MRI-Operations-Manual-V7.pdf>

**3.3 Progressive Neocortical Damage in Epilepsy (PNDE)**

Number of volunteers: 49

Reference for dataset: *Liu, R. S. N. et al. A Longitudinal Quantitative MRI Study of Community-Based Patients with Chronic Epilepsy and Newly Diagnosed Seizures: Methodology and Preliminary Findings. NeuroImage 14, 231–243 (2001).*

MR-acquisition: Each subject had a baseline and repeat T1-weighted inversion-recovery prepared volume acquisition (Fast InversionRecovery [prepared] Spoiled Gradient Recalled [IR–SPGR]:TI/TR/TE, 450/15/4.2 (ms), flip angle 25 degrees;124 1.5mm-thick coronal slices; matrix, 256, 192 voxels,24, 18cm field of view; scan time, 7 minutes). This covered the whole brain with voxel sizes of 0.9375, 0.9375,1.5mm.

**4. Propensity Score Matching**

We used propensity score matching to match healthy controls to the NORSE cohort based on age and sex. Nearest-neighbour matching was performed in a 3:1 ratio using the MatchIt package in R version 4.4.1 (R Foundation for Statistical Computing). This resulted in successful matching of 33 NORSE cases with 99 healthy controls.

**Table 6:** *NORSE and healthy control demographics after matching*

|  | NORSE  (n = 33) | Healthy controls  (n = 99) | Standard mean difference | P value |
| --- | --- | --- | --- | --- |
| **Sex** |  |  |  |  |
| **Female** | 16 (48%) | 49 (49%) | 0.02 | 1.00 |
| **Male** | 17 (52%) | 50 (51%) |  |  |
| **Age at baseline scan *(years)*** | 47 ± 16 | 50 ± 20 | 0.17 | 0.39 |

**5. Two example NORSE cases**

**5.1 Case 1**

A previously healthy man in his late 30-ties was admitted to a regional Swiss hospital with a four-day history of febrile illness (maximum temperature: 40.5 °C), fatigue, and progressive somnolence. During emergency transport, he experienced a generalized tonic-clonic seizure, successfully aborted with intravenous midazolam. He was intubated upon arrival.

Initial blood tests revealed a mildly elevated C-reactive protein (CRP: 20 mg/L). Cerebrospinal fluid (CSF) analysis showed 6 mononuclear cells and markedly elevated protein. Based on a working diagnosis of meningoencephalitis, empirical treatment with intravenous ceftriaxone, acyclovir, and dexamethasone was initiated. Head CT and brain MRI (performed on day 1) were unremarkable, and the initial EEG showed no abnormalities.

Seizure management began with levetiracetam and midazolam but required escalation due to breakthrough seizures during attempts to reduce antiseizure medication (ASM). Seizures presented as left facial myoclonus progressing to secondary generalization on the left side of the body. Additional treatments included valproic acid, sufentanil, phenytoin, thiopental, intermittent lorazepam and propofol, and high-dose corticosteroids over five days.

After one week of refractory seizures, the patient was transferred to a tertiary hospital. EEG confirmed super-refractory SE with clinical correlates. Follow-up MRIs showed right frontal cortical FLAIR hyperintensities, raising suspicion of focal cortical dysplasia. However, a brain biopsy revealed only non-specific inflammatory changes, and FDG-PET did not detect any metabolically active lesions.

Extensive diagnostic testing—including vasculitis screening, HIV and hepatitis serologies, viral metagenomics, onconeural antibodies, and autoimmune encephalitis panels—yielded negative results. The only consistent CSF abnormality was a marked elevation in Tau protein, with negative RT-QuIC. Tau elevation was interpreted as a consequence of ongoing SE.

Despite five cycles of plasmapheresis and multiple off-label interventions (including lidocaine, sevoflurane, high-dose magnesium, pregabalin, and ketogenic diet), the seizures remained refractory. After six weeks, the patient began to show signs of clinical improvement, including nonverbal communication. EEG gradually improved, though epileptic discharges persisted in the right frontal region.

Following nearly five months of hospitalization, the patient was transferred to a rehabilitation facility on a regimen of lorazepam, phenobarbital, piracetam, and pregabalin. However, his condition deteriorated during rehabilitation, requiring re-hospitalization due to increasing seizure frequency. After temporary stabilization, he returned to the rehabilitation center but soon developed a new episode of SE, which proved unresponsive to treatment. He subsequently died due to uncontrollable seizure activity.

**5.2 Case 2**

A woman in her late forties with no prior medical history aside from right-hand arthritis, treated with leflunomide, developed headache and fever without focal neurological deficits following a cosmetic eyelid lift. A few days later, she reported a sensation of heat and hearing disturbances, followed by progressive somnolence and stupor. She was initially admitted to a psychiatric hospital and subsequently transferred to a regional Swiss hospital for further evaluation.

Initial brain MRI was unremarkable. At presentation, her temperature was 38.1 °C. Lumbar puncture revealed 34 leukocytes, elevated protein, and elevated glucose. Empiric treatment with intravenous ceftriaxone, acyclovir, and dexamethasone was initiated for suspected meningoencephalitis.

While in the emergency department, she experienced her first seizure, presenting with myoclonus of the right mouth and right foot. The seizure was initially controlled with levetiracetam, but recurred shortly thereafter, necessitating continuous midazolam infusion and the addition of valproic acid. The first EEG showed bifrontal epileptiform activity.

She was transferred to a university hospital, where MRI revealed bilateral edematous changes in the basal ganglia. Continuous EEG confirmed ongoing SE, which persisted for several weeks. The diagnosis of new-onset refractory status epilepticus (NORSE) was made.

Extensive diagnostic work-up—including encephalitis panels, vasculitis and rheumatologic screening, a comprehensive viral panel, antineural antibodies, and RT-QuIC—was negative. Brain biopsy revealed reduced neuronal numbers, reactive astrogliosis and microgliosis and occasional paravasal macrophages.

Multiple immunotherapies—including corticosteroids, plasmapheresis, intravenous immunoglobulin (IVIG), and cyclophosphamide—were administered without clinical improvement. On the ICU, she received a broad spectrum of antiseizure medications (ASM), including midazolam, thiopental, ketamine, propofol, magnesium, and lidocaine, without meaningful seizure control. Improvement was observed only after the addition of phenobarbital, clonazepam, and piracetam. SE stopped although focal seizures with oral myoclonus persisted.

Her condition gradually improved over the following weeks. Complete seizure control was achieved initially. Once vigilance improved, she was transferred to a neurorehabilitation facility, where she was able to communicate using head nods and shakes to answer yes/no questions.

To date, no definitive aetiology has been established. Approximately six months after discharge, she experienced recurrent SE triggered by a urinary tract infection. Over the subsequent years, her condition slowly improved. She was able to communicate with brief sentences, walk a couple of meters with help and had moderate to severe residual cognitive deficits. More than one year after the initial NORSE she started having seizures with jerking in her face and arm and rare tonic-clonic seizures. At the time of this report, she is being treated with levetiracetam, clobazam, lamotrigine, and sultiam. A recent hospitalization was reported due to increased seizure frequency.

**6. Power and Sample Size Calculation**

We calculated the required sample size with a closed-form formula that is algebraically equivalent to analysing the change score with a two-sample t-test but yields identical power for a random-intercept linear mixed-effects model with two time points.

We based the estimated rate of change during NORSE on previous research. Previous reports observed a 16% decrease in total brain volume, 23% hippocampal atrophy, and a 23% reduction in the brain-ventricle ratio in individuals with peri-ictal MRI abnormalities, nonconvulsive seizures, and super-refractory SE. References:

Hocker, S., Nagarajan, E., Rabinstein, A. A., Hanson, D. & Britton, J. W. Progressive Brain Atrophy in Super-refractory Status Epilepticus. *JAMA Neurol* **73**, 1201–1207 (2016).

Vespa, P. M. *et al.* Nonconvulsive seizures after traumatic brain injury are associated with hippocampal atrophy. *Neurology* **75**, 792–798 (2010).

Varela, P. B. *et al.* Brain damage caused by status epilepticus: A prospective MRI study. *Epilepsy Behav.* **161**, 110081 (2024).

Thus, we assumed a minimum mean 16% change of grey matter volume between scans during NORSE. In healthy controls, we assumed a maximum mean 1% change of grey matter volume across the same interval, reference:

Ge, R. *et al.* Normative modelling of brain morphometry across the lifespan with CentileBrain: algorithm benchmarking and model optimisation. *Lancet Digit. Heal.* **6**, e211–e221 (2024).

Assuming a true absolute difference in change of 15% (16% vs. 1%), a measurement standard deviation (σ) of 0.20 and a within-subject correlation (ρ) of 0.50, the analysis yielded **n = 14** per group for 80 % power at a two-sided α = 0.05.
